# Adaptive resistance to SHP2-based vertical RAS-pathway inhibition in pancreatic cancer involves multifaceted routes towards dedifferentiation

**DOI:** 10.1101/2025.09.09.25335395

**Authors:** Xun Chen, Steffen Johannes Keller, Tonmoy Das, Philipp Hafner, Asma Y. Alrawashdeh, Huda Jumaa, Ting Chen, Solène Besson, Johana Norona, Mara Schneider, Hanna Scheffold, Kerstin Meyer, Stephanie Mewes, Silke Hempel, Dominik von Elverfeldt, Wilfried Reichardt, Melanie Boerries, Stefan Fichtner-Feigl, Geoffroy Andrieux, Dietrich Alexander Ruess

**Author notes:** contributed equally (Xun Chen, Steffen Johannes Keller).

## Abstract

**Background:** Pancreatic ductal adenocarcinoma (PDAC) critically depends on oncogenic KRAS signaling. The Src-homology 2 domain-containing phosphatase 2 (SHP2) is essential for full KRAS activity. Allosteric SHP2 inhibition in vertical combination with mitogen-activated protein kinase (MAPK) or RAS inhibition demonstrates synergistic efficacy, delays the onset of resistance, and is currently being evaluated in clinical trials.

**Methods:** Employing a comprehensive set of models, including endogenous murine tumors, murine and human cell lines, and patient-derived organoids representing the full spectrum of PDAC molecular subtypes, we here explore the characteristics and mechanisms of adaptive resistance to dual SHP2/mitogen-activated protein kinase kinase (MEK1/2) and dual SHP2/KRAS^G12D^ inhibition.

**Results:** We observe potent anti-tumor efficacy of SHP2-based vertical RAS-pathway inhibition across all molecular PDAC subtypes. However, resistance eventually emerges and is commonly associated with tumor cell dedifferentiation. This dedifferentiation is marked by features of epithelial-to-mesenchymal transition (EMT), a shift in transcriptional molecular PDAC subtype, and cancer cell state transitions. Resistance involves increased RAS expression and/or context-dependent activation of various compensatory signaling pathways, including PI3K-AKT, JAK-STAT, or Wnt/β-catenin, Hippo, Rap1 and TGFβ.

**Conclusion:** These findings suggest that resistance to SHP2-based vertical RAS-pathway inhibition follows converging, yet molecularly heterogeneous, compensatory signaling trajectories. Consequently, precision medicine approaches that target specific signaling alterations or exploit the vulnerabilities of dedifferentiated cell states are necessary to overcome resistance and reinforce vertical RAS-pathway inhibition for effective pancreatic ductal adenocarcinoma (PDAC) treatment.

## Introduction

Pancreatic ductal adenocarcinoma (PDAC) remains one of the most aggressive malignancies, with mortality rates nearly matching incidence. Despite its comparatively low prevalence, PDAC is projected to become the second leading cause of cancer-related deaths in the United States in the near future ^1,2^. Recent therapeutic advances in the form of multiagent chemotherapy regimens, first precision medicine options, and interdisciplinary sequential therapy approaches have improved 5-year survival rates to approximately 13% ^3,4^. Surgical resection is required if curation is the goal, but only about 20-30% of cases present with or reach a resectable stage, and recurrence is common even after standard adjuvant chemotherapy ^2^.

This emphasizes the critical need to advance and apply innovative treatment approaches that can improve outcomes for a larger patient population. In recent years, promising preclinical and early clinical results have raised hope and expanded the therapeutic landscape of pancreatic cancer. These results include approaches such as personalized neoantigen-mRNA vaccines and RAS-targeting small molecules ^5–7^.

Oncogenic *KRAS* mutations are found in more than 90% of PDAC tumors, and *KRAS* wild-type PDAC frequently harbor genetic alterations in other components of the RAS signaling pathway ^8^. This underscores a profound dependence of PDAC on RAS-mediated signaling, promoting diverse cellular processes including proliferation, differentiation, survival, and metabolic adaptation ^9^.

Based on transcriptional expression profiles, PDAC can be subclassified into a clinically relevant spectrum of molecular subtypes ^10–14^. Patients with tumors characterized by a more dedifferentiated, more mesenchymal tumor cell pattern (basal-like subtype) have a poorer prognosis compared to those with tumors exhibiting a more differentiated, epithelial expression profile (classical subtype). While early studies suggested that differentiated epithelial tumor cells are more dependent on KRAS signaling ^15,16^, more recent findings have shown that increased gene dosage of mutant KRAS is associated with a more dedifferentiated and aggressive phenotype ^11,17,18^. These insights further support the concept that RAS signaling plays a critical role across the entire PDAC molecular subtype spectrum.

Given this dependency, the recent development of direct KRAS and RAS inhibitors has already made a tremendous impact ^6,7^, and will very likely improve patient outcomes substantially. However, as with MEK or ERK inhibitors, therapeutic resistance emerges rapidly and combination strategies are needed to achieve durable responses ^19–21^.

Previous studies have demonstrated that vertical RAS-pathway inhibition by dual targeting of SHP2 phosphatase and MEK1/2 or ERK1/2 overcomes RTK-reactivation dependent resistance mechanisms and leads to synergistic growth inhibition of PDAC tumors ^22–25^. Accordingly, co-targeting SHP2 is also a promising strategy in combination with (K)RAS directed agents ^26,27^. Several clinical trials are currently evaluating allosteric SHP2 inhibitors in combination with MEK1/2, ERK1/2, or RAS inhibitors (e.g. *NCT04916236, NCT04699188, NCT06416410*).

However, since therapy evasion has to be anticipated even with SHP2-based vertical RAS-pathway inhibition ^27^, we here aimed to elucidate the characteristics and underlying mechanisms of adaptive resistance in PDAC in response to dual SHP2/MEK1/2 or SHP2/KRAS^G12D^ inhibition. Our findings indicate that a variety of signaling pathways may contribute to dedifferentiation, EMT and transcriptional subtype transition, which occurs as a common pattern of adaptive resistance. These insights provide a rational basis for the design of future combination therapies tailored to prevent or overcome resistance and enhance the durability of RAS-pathway-targeted treatments.

## Results

### Development of adaptive resistance to dual SHP2/MEK inhibition *in vivo*

To evaluate the evolving patterns of resistance to vertical RAS-pathway inhibition via combined SHP2 plus MEK inhibition in PDAC *in vivo*, we utilized a gold-standard endogenous genetic murine model based on Cre-mediated expression of mutant Kras^G12D^ from the endogenous *Kras* locus and homozygous deletion of the tumor suppressor gene *Trp53* during embryonic development (*Ptf1a^Cre-ex^*^1^), in the following referred to as KPC (**Figure 1A**). In this model, multifocal, multiclonal and heterogeneous tumors develop *in situ*, progressively transforming the whole pancreas. In parallel, a complex tumor microenvironment evolves, most faithfully recapitulating the human condition. KPC mice were monitored longitudinally by weekly MRI, both before and during the therapeutic trial. In contrast to our previous work ^22^, we replaced the protein tyrosine phosphatase (PTP) catalytic site inhibitor GS493, which is limited by poor pharmacokinetics and specificity, with the orally bioavailable, allosteric SHP2 inhibitor SHP099. The well-documented robust short-term response to Trametinib (MEK inhibitor), measured by MRI-based pancreas volume reduction (**Figure 1B**), (relative) pancreatic weight (**Figure 1C**) and Ki67 immunohistofluorescence (**Figure 1D**), was confirmed. This short-term effect was slightly further enhanced by the addition of SHP099 in the combination therapy arm. Of note, and in contrast to previous observations with GS493 ^22^, already as monotherapy SHP099 demonstrated statistically significant short-term therapeutic effect (**Figure 1C and Figure 1D**).

**Figure 1.**
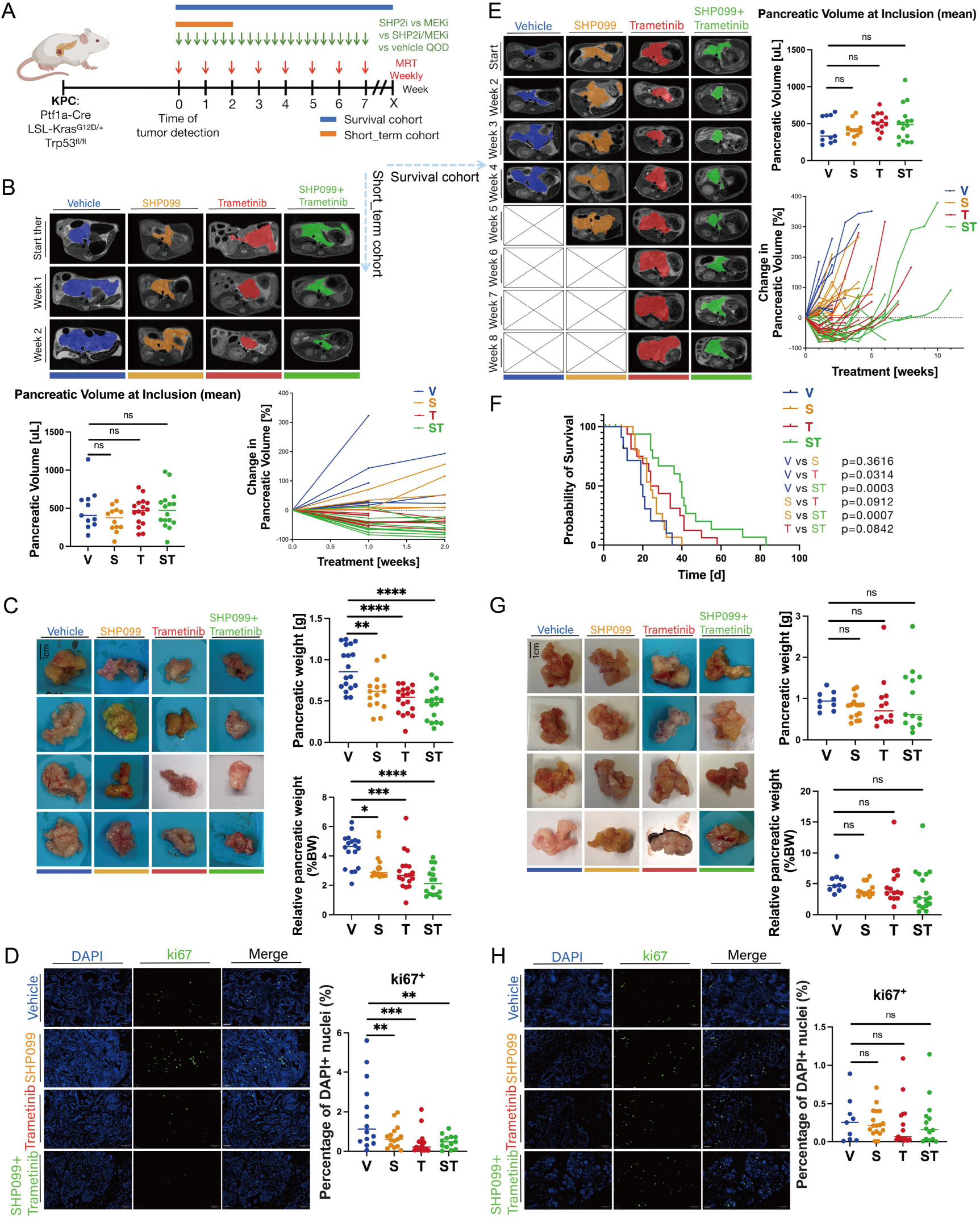
Substantial but short-lived *in vivo* response of autochthonous PDAC to vertical RAS-pathway interference by orally administered dual SHP2/MEK inhibition. (**A**) Experimental work-flow with the autochthonous KPC mouse model. (**B-D**) short term (2 week) treatment cohorts: (**B**) MRI monitoring of pancreatic volume changes over time. Representative sections of exemplary mice taken at indicated time intervals after administration of four different conditions as indicated (blue, orange, red and green outlines represent pancreatic tumors). Pancreatic volumes at inclusion were rather comparable across conditions. V: Vehicle; S: SHP099; T: Trametinib; ST: SHP099+Trametinib. ns: not significant. (**C**) Following short-term treatment, the whole pancreas was harvested and weighed. Pancreatic weight was put in relation to whole body weight. Four representative photographs of pancreas specimens of each cohort are displayed. (**D**) Ki67-immunohistofluorescence of KPC pancreata from the short-term treatment cohorts. Representative images are shown. Scale bars: 50 μm. (**E-H**) long term (survival) treatment cohorts: (**E**) MRI monitoring of pancreatic volume changes over time. Representative sections of exemplary mice taken at indicated time intervals after administration of four different conditions as indicated (blue, orange, red and green outlines represent pancreatic tumors. (**F**) Kaplan-Meier overall survival analysis of all mice enrolled in the different survival trial arms. Vehicle (blue): n=11; SHP099 (orange): n=17; Trametinib (red): n=16; SHP099+Trametinib (green): n=18. (**G**) After survival endpoint, pancreatic tissue was harvested and weighed and put in relation with whole body weight. Four representative photographs of pancreas specimens of each cohort are displayed; (**H**) Ki67-immunohistofluorescence of KPC pancreata from the survival treatment cohorts. Representative images are shown. Scale bars: 50 μm. Statistical significance was determined via one-way ANOVA in panels B, C, D, E, G, H, with comparisons made against corresponding vehicle controls. ****: p<0.0001; ***: p<0.001; **: p<0.01; ^ns^: p>0.05.

Additional KPC mice were then subjected to continuous, long-term treatment until reaching a humane survival endpoint. As previously described, monitoring via weekly MRI revealed a transient reduction in tumor volume with Trametinib for 2-4 weeks, followed by tumor regrowth and uncontrolled progression (**Figure 1E**). In contrast, the addition of SHP099 to Trametinib enabled more durable therapy effects and conferred a more substantial survival benefit in this highly aggressive PDAC model (**Figure 1F**). Nevertheless, while dual SHP099/Trametinib inhibition effectively delayed disease progression, it ultimately failed to prevent resistance development and tumor regrowth (**Figure 1E**). Consequently, at the time of sacrifice, post-treatment relative pancreatic weights did not differ significantly among the four treatment groups (**Figure 1G**), and in contrast to the short-term treatment cohort, tumors in the long-term survival treatment group showed evidence of regained proliferation (**Figure 1H**), indicating therapy evasion.

Together, these findings highlight a striking primary sensitivity of endogenous PDAC to combined SHP2/MEK inhibition. This vertical RAS-pathway targeting approach effectively mitigates resistance dynamics and prolongs overall survival in an aggressive tumor model. However, these therapeutic benefits are ultimately transient, as tumors eventually adapt and escape, underscoring the urgent need to overcome adaptive resistance mechanisms.

### Dual SHP2/MEK inhibition: Response and resistance patterns stratified by molecular subtype

To further investigate subtype-specific patterns of adaptive resistance to dual SHP2/MEK inhibition in a more controlled setting we conducted *in vitro* analyses using KPC-derived cell lines with defined phenotypic and transcriptional profiles. We established several murine PDAC cell lines (one per KPC mouse), and after achieving stable passaging, characterized their molecular subtype through morphological assessment, western blotting for subtype-associated markers, and RNA sequencing followed by single-sample gene set enrichment analysis (ssGSEA) using published PDAC subtype gene sets^10–14^ (**Figure 2A**, **Supplemental Figure 1A**). Integration of this data revealed that our collection of KPC cell lines represents a continuous spectrum of subtypes, ranging from strongly basal-like/mesenchymal over intermediate to clearly classical/epithelial, thereby reflecting intertumoral heterogeneity observed in patients (**Figure 2A**). We then selected two basal-like (#495 and #382) and two classical (#215 and #162) KPC cell lines for comparative therapy response evaluation and induction of adaptive resistance.

**Figure 2.**
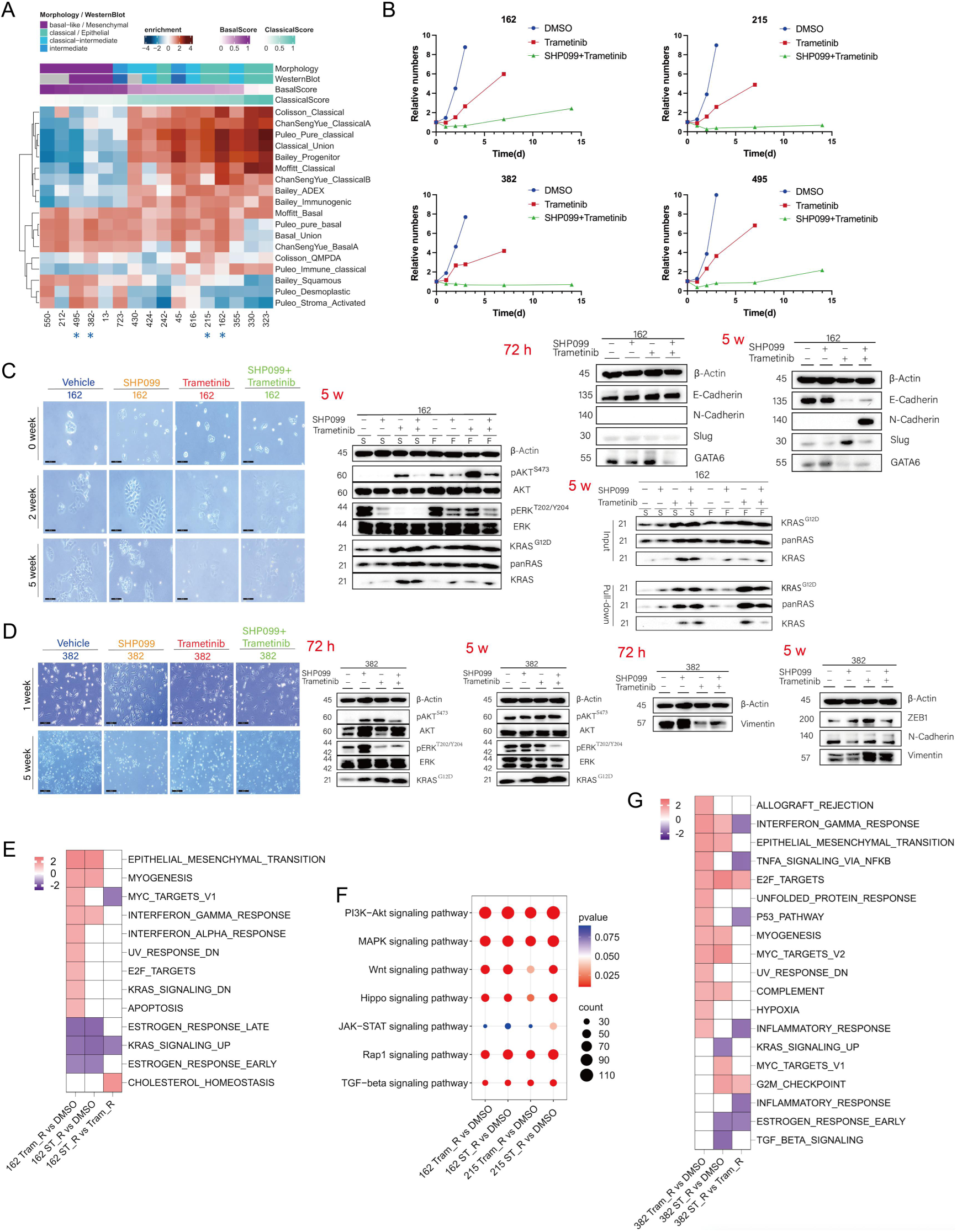
Dual SHP2/MEK inhibition: Response and resistance patterns stratified by molecular subtype *in vitro*. (**A**) Unsupervised clustering of transcriptional subtype analysis (ssGSEA) of untreated murine PDAC cell lines derived from KPC tumors. The Basal Score and Classical Score were defined by us as overarching categories, consolidating published subtype signatures corresponding to the basal-like and classical phenotypes, respectively (primary author and suggested subtype terminology for single gene sets are indicated on the right). Light microscopic morphology and PDAC subtype related protein expression levels by Western Blot are indicated. The four cell lines selected for further functional analyses are indicated by blue stars (*). (**B**) Proliferation experiments were performed over 14 days *in vitro* using different treatment arms, and cell counts were performed as indicated. (**C+D**) Light microscopic appearance and immunoblot assays with the indicated antibodies of the short-term treated (72 h) and drug-resistant (5 w) KPC “classical” cell line 162 in C and KPC “basal-like” cell line 382 in D. Scale bars: 200 μm. β-Actin served as the loading control. S: FBS starvation the day before lysing the cells; F: FBS starvation the day before + supplying FBS 10 minutes before lysing the cells. Pulldown of RAS-GTP was achieved using RAF-RBD agarose beads. (**E**) Normalized enrichment scores (NES) from the Hallmark gene sets comparing resistant KPC cell lines derived from parental line 162 “classical” (162 Tram_R and 162 ST_R) with a DMSO-treated control. Only gene sets with adj. p < 0.05 are indicated. (**F**) Pathway analysis of key signaling pathways altered in resistant PDAC cell lines (from parental 162 and 215, “classical”). Dot size indicates gene count; color reflects statistical significance (adj. p-value). (**G**) NES from the Hallmark gene set comparing resistant KPC cell lines derived from parental line 382 “basal-like” (382 Tram_R and 382 ST_R) with a DMSO-treated control. Only gene sets with adj. p < 0.05 are indicated.

First, we again used and compared MEK and dual SHP2/MEK inhibition. We measured cell proliferation over a period of two weeks of therapy. Dual SHP2/MEK inhibition was substantially superior to MEK inhibition alone for all four cell lines, to which all lines quickly adapted (**Figure 2B**). Subsequently, we induced treatment resistance by continuously exposing these four cell lines to fixed drug concentrations of SHP099 (S), Trametinib (T) and dual SHP099/Trametinib (S/T) until they resumed proliferation; the workflow is depicted in **Supplemental Figure 1B**.

With the two classical lines (#215 and #162) we observed a pronounced morphological shift toward a mesenchymal phenotype of the drug-resistant populations, especially with S/T treatment (**Figure 2C**, **Supplemental Figure 1C**). Look into signaling pathway alterations, we found that ERK phosphorylation remained suppressed, particularly with S/T, and AKT phosphorylation increased, most prominently in #215. (**Figure 2C**, **Supplemental Figure 1C**). Simultaneously, KRAS and KRAS^G12D^ protein levels and GTP-loading were elevated in T and S/T resistant cells. Screens for expression of EMT related proteins revealed a downregulation of E-Cadherin and strong upregulation of N-Cadherin expression in drug-resistant cell lines, the latter being completely absent in the treatment naïve parental classical cell lines #215 and #162. In contrast, the two basal cell lines (#382 and #495) did not exhibit significant changes in microscopic cell morphology and retained their features characteristic of a mesenchymal cell phenotype (**Figure 2D**, **Supplemental Figure 1D**) in the resistant state. However, immunoblot and signaling pathway analysis revealed features in common with the two classical cell lines, with maintained inhibition of ERK phosphorylation, most prominently in the context of S/T resistance, and upregulation of KRAS^G12D^ expression in response to T and S/T. But compensatory AKT or STAT3 activation was less obvious in those lines. Protein levels also suggested a shift further towards the mesenchymal end of the EMT spectrum, with epithelial (classical) markers like E-Cadherin and GATA6 remaining completely absent, and mesenchymal markers like Vimentin (#382) and Slug (#382 and #495) demonstrating increased expression in T and S/T resistant populations (**Figure 2D**, **Supplementary Figure 1D**).

We then extended these analyses to KPC cell lines that were obtained from tumors that had been treated and become resistant *in vivo*. Cell lines isolated from T and S/T *in vivo* long-term treated tumors had mesenchymal morphology, those from vehicle and S treated tumors epithelial phenotype (**Supplemental Figure 1E**). We continued long-term treatment *ex vivo in vitro*, which did not change morphology. We again performed immunoblot analysis and observed sustained inhibition of ERK phosphorylation most prominently in the context of S/T treatment, and compensatory AKT phosphorylation already with S, but more prominently with T and S/T. Again, upregulation of KRAS expression and an increase in GTP-loading of panRAS and KRAS^G12D^ was detected with T and S/T treatment. Additionally, there was a significant elevation in STAT3 phosphorylation in cell lines exhibiting resistance to T treatment. Moreover, immunoblot findings demonstrated a significant downregulation of E-Cadherin in T and S/T and a notable upregulation of N-Cadherin in S/T drug-resistant cell lines (**Supplemental Figure 1E**). To assess the reversibility of resistance, we withdrew treatment from our *ex vivo in vitro* KPC cell lines for three weeks. In the S/T-resistant cells, ERK phosphorylation recovered and AKT activation subsided, suggesting a plastic, reversible phenotype. In contrast, our T-resistant cells maintained high AKT and STAT3 activity. These results confirmed two possible avenues for resistance to targeted therapy, i.e. cellular plasticity vs. a fixed resistant state, the latter possibly caused by genetic alterations acquired during *in vivo* clonal selection (**Supplemental Figure 1E**).

We then performed RNA sequencing followed by GSEA, which confirmed that EMT-related gene sets were most dominantly and significantly enriched in T and dual S/T drug-resistant cell lines across our cell line panel, irrespective of molecular subtype (**Figure 2E**, **G**, **Supplemental Figure 2A**, **B**, **Supplemental Figure 3A**). Concurrently, we observed compensatory enrichment of PI3K-AKT, MAPK, Wnt, Hippo, JAK-STAT, Rap1 and TGF-beta pathway related gene sets in both, basal-like and classical, T and dual S/T resistant cell lines (**Figure 2F**, **Supplemental Figure 2C**, **Supplemental Figure 3B**).

These results indicated that both, basal-like and classical KPC-derived PDAC cell lines display an initially high sensitivity to dual SHP2/MEK inhibition. Yet eventually, adaptive resistance emerges across subtypes. Resistance is associated with largely sustained suppression of MAPK-pathway, elevated RAS expression and GTP-loading, and compensatory activation of alternate signaling pathways, most prominently PI3K-AKT, and a shift toward the mesenchymal extreme of the EMT spectrum. Classical epithelial lines change morphology and basal-like, already mesenchymal lines further increase expression of EMT-related proteins.

### Resistance patterns evolving with dual SHP2/KRAS^G12D^ inhibition

Given the broad efficacy of SHP2/MEK inhibition across the molecular subtype spectrum of murine PDAC and the common EMT-pattern in the context of resistance we extended our analyses to KRAS^G12D^ and dual SHP2/KRAS^G12D^ inhibition. Again, we established resistant cell lines from the same treatment naïve cell lines as above (classical: #162 and #215; basal-like: #382 and #495) by treating them with the KRAS^G12D^ off-state inhibitor MRTX1133, either alone or in combination with SHP099 (workflow see **Supplemental Figure 1B**). We then again assessed morphology, signaling alterations, EMT marker expression and transcriptional changes.

Proliferation assays over the first 14 days revealed that basal-like cell lines responded more prominently to MRTX1133 monotherapy than classical lines. Notably, dual SHP099/MRTX1133 treatment resulted in complete proliferation arrest in all four cell lines throughout this treatment duration (**Figure 3A**), indicating potent synergistic activity.

**Figure 3.**
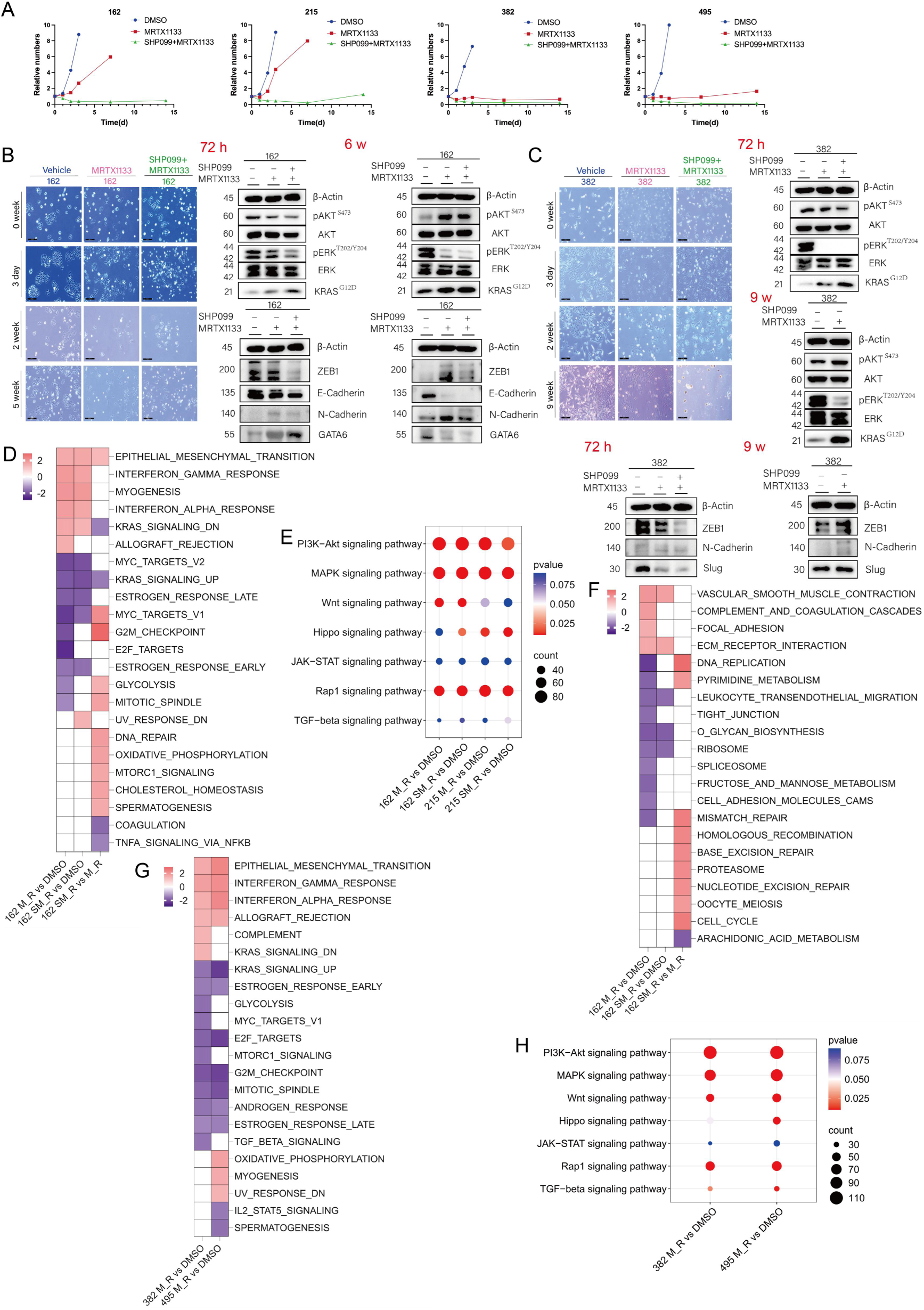
Dual SHP2/KRAS inhibition: Response and resistance patterns stratified by molecular subtype *in vitro*. (**A**) Proliferation experiments were performed over 14 days in *vitro* using different treatment arms, and cell counts were performed at different time points, as indicated. (**B+C**) Light microscopic appearance and immunoblot assays with the indicated antibodies of short-term treated (72 h) and drug-resistant KPC “classical” cell line 162 (6 w) in B and KPC “basal-like” cell line 382 (9 w) in C. Scale bars: 200 μm. β-Actin served as the loading control. (**D**) NES from the Hallmark gene sets comparing resistant KPC cell lines derived from parental line 162 “classical” (162 M_R and 162 SM_R) with a DMSO-treated control. Only gene sets with adj. p < 0.05 are indicated. (M: MRTX1133; SM: SHP099+MRTX1133). (**E**) Pathway analysis of key signaling pathways altered in resistant PDAC cell lines (from parental 162 and 215, “classical”). Dot size indicates gene count; color reflects statistical significance (adjusted p-value). (**F**) KEGG gene set enrichment of genes in resistant KPC cell lines derived from parental lines 162 “basal” (162 M_R and 162 SM_R). Only gene sets with adj. p < 0.05 are indicated. (**G**) NES from the Hallmark gene set comparing resistant KPC cell lines derived from parental lines 382 and 495, “basal-like” (382 M_R and 495 M_R) with DMSO-treated controls. Only gene sets with adj. p < 0.05 are indicated. (M: MRTX1133). (**H**) Pathway analysis of key signaling pathways altered in resistant KPC cell lines derived from parental lines 382 and 495, “basal-like”. Dot size indicates gene count; color reflects statistical significance (adj. p-value).

In classical KPC cell lines (#162 and #215), acquired resistance was again associated with a morphological shift toward a mesenchymal phenotype (**Figure 3B**, **Supplementary Figure 4A**). Again, ERK phosphorylation remained suppressed, whereas AKT and STAT3 phosphorylation was increased, particularly in the context of dual SHP2/KRAS^G12D^ inhibition resistance. Additionally, expression of pan-RAS, total KRAS, and KRAS^G12D^ was upregulated. EMT marker profiling revealed E-Cadherin downregulation and N-Cadherin upregulation, evident already with MRTX1133 monotherapy but more pronounced with the dual treatment.

MRTX1133 resistant basal-like cell lines (#382 and #495) retained a mesenchymal cell phenotype (**Figure 3C**, **Supplementary Figure 4B**). ERK phosphorylation remained inhibited. However, cell line-specific signaling alterations were observed: line #382 strongly upregulated expression of the KRAS oncogene and demonstrated slight increase in AKT activation, and the other (#495) appeared to virtually have lost KRAS expression and had lower AKT phosphorylation levels. EMT marker expression also varied and lacked consistency (**Figure 3C**, **Supplementary Figure 4B**), likely due to clonal selection of a minor population within the parental cell line, with more epithelial characteristics. Importantly, due to the profound and sustained proliferation inhibition induced by dual SHP099/MRTX1133, we were unable to generate basal-like cell lines resistant to this combination, underscoring the therapeutic potential of this strategy.

Transcriptomic analysis further supported these findings. In classical cell lines, EMT-related gene sets were significantly enriched (**Figure 3D**, **F**, **Supplementary Figure 4C, D**). Enrichment of PI3K-AKT, MAPK, Wnt, Hippo, and Rap1 signaling pathways was also again observed (**Figure 3E**). Very similarly, in basal-like cell lines, transcriptomic profiling also showed strong enrichment of EMT-related gene sets in response to KRAS^G12D^ inhibition (**Figure 3G**, **Supplementary Figure 4E**). Parallel enrichment of PI3K-AKT, MAPK, Wnt, Hippo, Rap1, and TGFβ signaling pathway genes was again observed (**Figure 3H**).

Taken together, these data indicated that treatment naïve basal-like cell lines exhibit particular sensitivity already to KRAS^G12D^ monotherapy, and are very effectively controlled by dual SHP2/KRAS^G12D^ inhibition. With some limitations in the protein-level EMT-analysis for the basal-like cell lines due to our oligoclonal parental cell lines generated by the outgrowth-method, patterns of resistance to MRTX1133 and dual SHP099/MRTX1133 across subtypes, regarding mesenchymal transition, (K)RAS expression as well as associated compensatory signaling alterations mirror those observed with SHP2/MEK inhibition. These findings indicate a convergent resistance mechanism induced by vertical RAS-pathway inhibition involving SHP2.

### Subtype transitions and cell state dynamics in vertical RAS-inhibition resistant KPC cell lines

Given the recurring emergence of an EMT phenotype, we hypothesized that vertical RAS pathway inhibition would induce molecular subtype transitions. To test this, we expanded our RNA-seq analyses to include all KPC cell lines introduced in **Figure 2A**, following short-term (48h) treatment with S, T, or S/T, and generated additional treatment resistant lines (#323: Tram_R and ST_R, and #330: Tram_R and ST_R). We also included transcriptomes from KPC cell lines with *in vivo ex vivo* continued treatment and their respective *in vitro* discontinuation counterparts (#212 Tram_cont, #550 Tram_cont, #45 ST_cont, #430 ST_cont, each vs. DMSO). We integrated all transcriptomic datasets and performed ssGSEA on single samples using established PDAC subtype gene sets. To facilitate integrative analyses, we defined two overarching categories – basal union and classical union – by consolidating published subtype signatures corresponding to the basal-like and classical phenotypes, respectively. Enrichment scores were averaged across replicates. Samples were then ranked according to the Basal (union) score (from high to low) (**Figure 4A**). While short-term treated samples largely retained subtype profiles resembling their DMSO-treated parental counterparts, treatment-resistant samples – particularly those derived from originally classical cell lines exposed to dual SHP2/MEK or dual SHP2/KRAS^G12D^ inhibition – displayed a marked shift along the subtype continuum toward a basal-like phenotype. In contrast, cell lines with a basal-like identity at baseline in part demonstrated somewhat decreased basal-like scores and increased classical-scores, but maintained their subtype following resistance development (**Figure 4A**).

**Figure 4.**
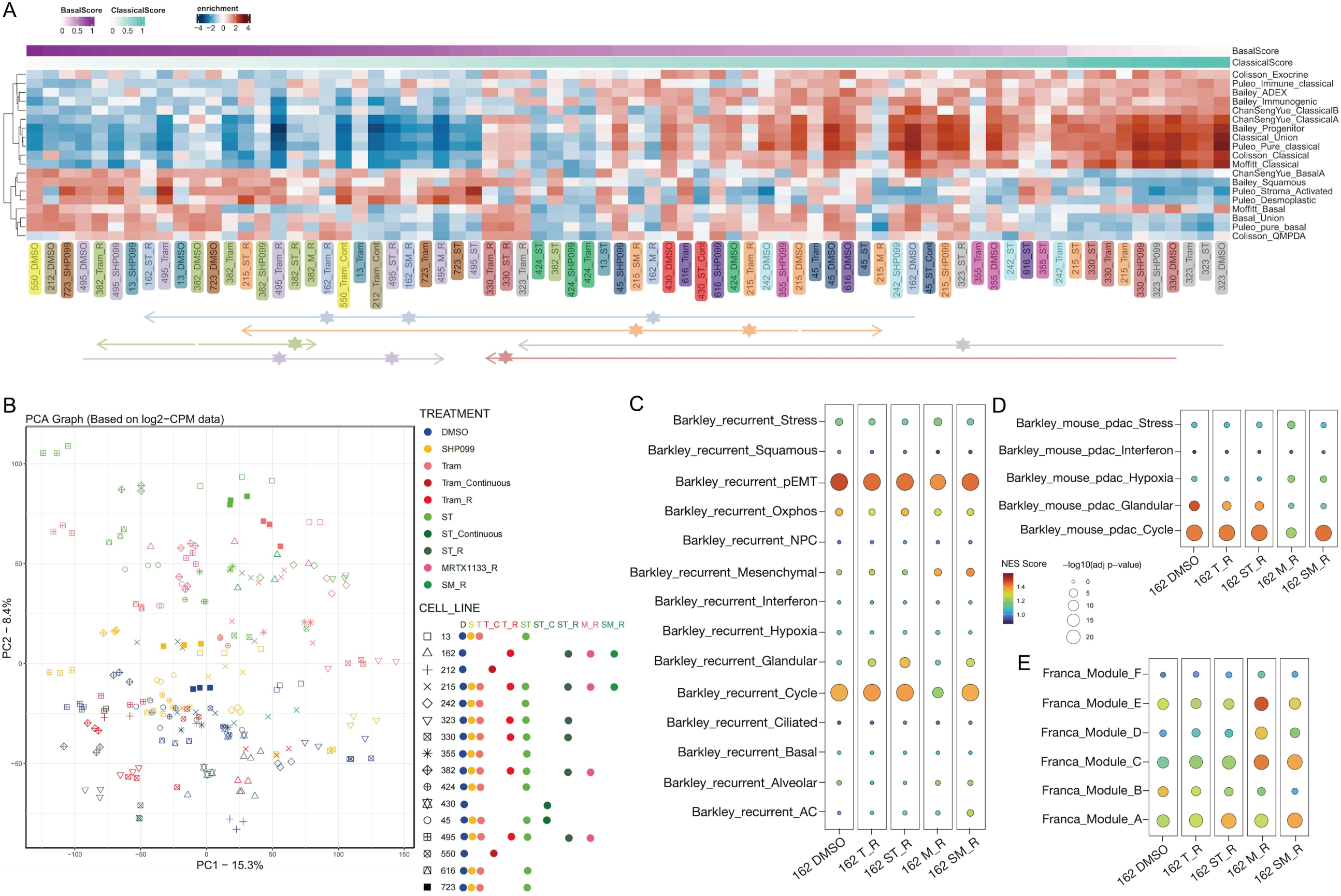
Transcriptional subtype plasticity and cell state dynamics in adaptive resistance to vertical RAS pathway inhibition in KPC cell lines. (**A**) Unsupervised clustering of transcriptional subtype analysis (ssGSEA) of murine PDAC cell lines derived from KPC tumors. Methodology as in Fig. 2A. Here, an expanded panel of long-term treated, adaptive resistant cell lines and their parental counterparts (DMSO treated) were included. Samples are grouped and color-coded according to parental-resistant counterparts. Labels of the same color indicate different states of the same cell line. Each arrow connects different treatment conditions of the same cell line. Arrows with identical colors indicate subtype transitions within the same cell line across treatments, reflecting therapy-induced transcriptional reprogramming. “Continuous” indicating ex vivo in vitro continuation of therapy. (**B**) Principal component analysis (PCA) of log₂-transformed CPM expression data from all samples across cell lines and treatments, illustrating transcriptional divergence among treatment groups. All samples from A consists of 3 individual triplicates, which were included in the PCA analysis as shown. Treatment groups are color-coded and cell lines represented by distinct shapes. (**C**) Single sample enrichment analysis (ssGSEA) using the catalog of gene modules whose expression defines recurrent cancer cell states described by Barkley et al. Dot plot showing enrichment of the recurrent cancer cell state gene modules and the murine PDAC cell state gene modules across multiple treatment conditions in the “classical” cell lines 162. NES and adj. p-values are visualized by color and dot size, respectively. (**D**) Mouse PDAC gene module analysis using the Barkley et al. gene sets. Dot plot showing enrichment of murine PDAC subtype gene modules across multiple treatment conditions in the “classical” 162 cell line. NES and adj. p-values are visualized by color and dot size, respectively. (E) Resistance continuum gene module enrichment analysis based on gene sets described by Franca et al. Dot plot illustrating enrichment of Modules A (earliest resistance stage/state) to F (latest resistance stage/state) across multiple treatment conditions in the “classical” 162 cell line. NES and adj. p-values are visualized by color and dot size, respectively.

Principal Component Analysis (PCA) of RNAseq data from all KPC cell lines revealed distinct patterns of transcriptional trajectories from DMSO-treated parental controls. Short term treatment with SHP2, MEK and combined SHP2/MEK inhibition progressively increased the distance from parental samples along a largely uniform direction across molecular subtypes. In contrast, MEK– and SHP2/MEK-resistant cell lines clustered in an almost opposite direction, but less distant to parental profiles than their short-term treated counterparts. Resistance to KRAS^G12D^ and SHP2/ KRAS^G12D^ inhibition however followed a PCA-trajectory more similar to short-term treatment patterns. Importantly, treatment-resistant populations that had undergone a classical-to-basal-like shift clustered distinctly from treatment-naïve basal-like parental lines.

These patterns support the notion that short-term treatment (esp. dual inhibition) induces marked transcriptional reprogramming, which is reversed during the process of adaptive resistance. They also point to differences between adaptive responses to Trametinib– and MRTX1133-based therapies, despite phenotypic similarities we observed before. And finally, they confirm that mesenchymal-like states emerging under therapeutic pressure are transcriptionally distinct from pre-existing basal-like phenotypes. This aligns with our earlier observations of their markedly different therapeutic responses – namely, the pronounced sensitivity of treatment naïve basal-like compared to the therapy-resistant mesenchymal cell lines (**Figure 4B**).

Prompted by recent descriptions of cancer cell states by Barkley et al. ^28^ and the concept of a resistance continuum involving cell state transitions by Franca et al. ^29^, we next assessed whether these concepts applied to our models. We performed ssGSEA using the gene modules reported by Barkley et al. and Franca et al., across four KPC lines (classical: #162, #215; basal-like: #382, #495) with short-term treatment and resistant samples available.

Of note, Barkley’s partial EMT (pEMT) signature was strongly enriched across all conditions, irrespective of treatment, suggesting inherent cellular plasticity as a hallmark of PDAC (**Figure 4C**, **Supplementary Figures 5A**, **B**, **C**). In several resistant samples, we observed gene set enrichment patterns consistent with mesenchymal cell states, including reduced enrichment for Barkley’s Glandular cell state signatures in classical lines #162 and #215 (**Figure 4D**, **Supplementary Figure 5A**), and increased Barkley’s Mesenchymal signature enrichment in #162 (**Figure 4C**), and partially a further increase in the basal-like cell line #495 (**Supplementary Figure 5C**). In all four KPC lines, resistance acquisition was associated with progression along the resistance continuum defined by Franca et al., which comprises six successive transcriptional states (A–F). We observed increased enrichment of modules C (EMT), D (EMT, NFκB signaling, translation), and E (translation, hypoxia, OXPHOS, ROS, ferroptosis), not uniformly across all samples, but consistently in at least one resistant sample per line (**Figure 4 E**; **Supplementary Figures 5A**, **B**, **C**). We also looked into the (human) PDAC programs suggested by Hwang et al. ^30^ but did not detect meaningful associations or shifts.

Together with the EMT data, these findings suggest that prolonged vertical RAS-pathway inhibition, either through dual SHP2/MEK or SHP2/KRAS^G12D^ targeting, induces convergent, yet not necessarily identical, transcriptional reprogramming. This reprogramming appears to be characterized by a common subtype shift toward the basal-like end of the PDAC spectrum and a cell state transition toward non-glandular, mesenchymal phenotypes along a previously described resistance continuum. Crucially, this adaptive response demarcates treatment-induced mesenchymal states from pre-existing basal-like subtypes.

### EMT accompanies evolving resistance *in vivo*

Having identified EMT and dedifferentiation as dominant and recurrent patterns of resistance in controlled *in vitro* settings, we next examined whether these processes were similarly involved in the heterogeneous tumors from our murine therapeutic trial.

We first applied standard immunohistochemistry to evaluate epithelial marker loss in tumors from KPC mice treated long-term with Trametinib or dual SHP099/Trametinib until survival endpoint. Notably, E-Cadherin expression was significantly reduced in tumors exposed to MEK or dual SHP2/MEK inhibition (**Figure 5A**). With long-term MEK inhibitor (MEKi) treatment, ERK phosphorylation did remain significantly suppressed, but a partial rebound in mean pERK levels was observed. In contrast, the dual SHP2/MEK inhibition cohort demonstrated lower average pERK levels and stronger statistical significance for pathway suppression. In both treatment arms, compensatory AKT phosphorylation was markedly increased, consistent with our previous *in vitro* findings (**Figure 5A**).

**Figure 5.**
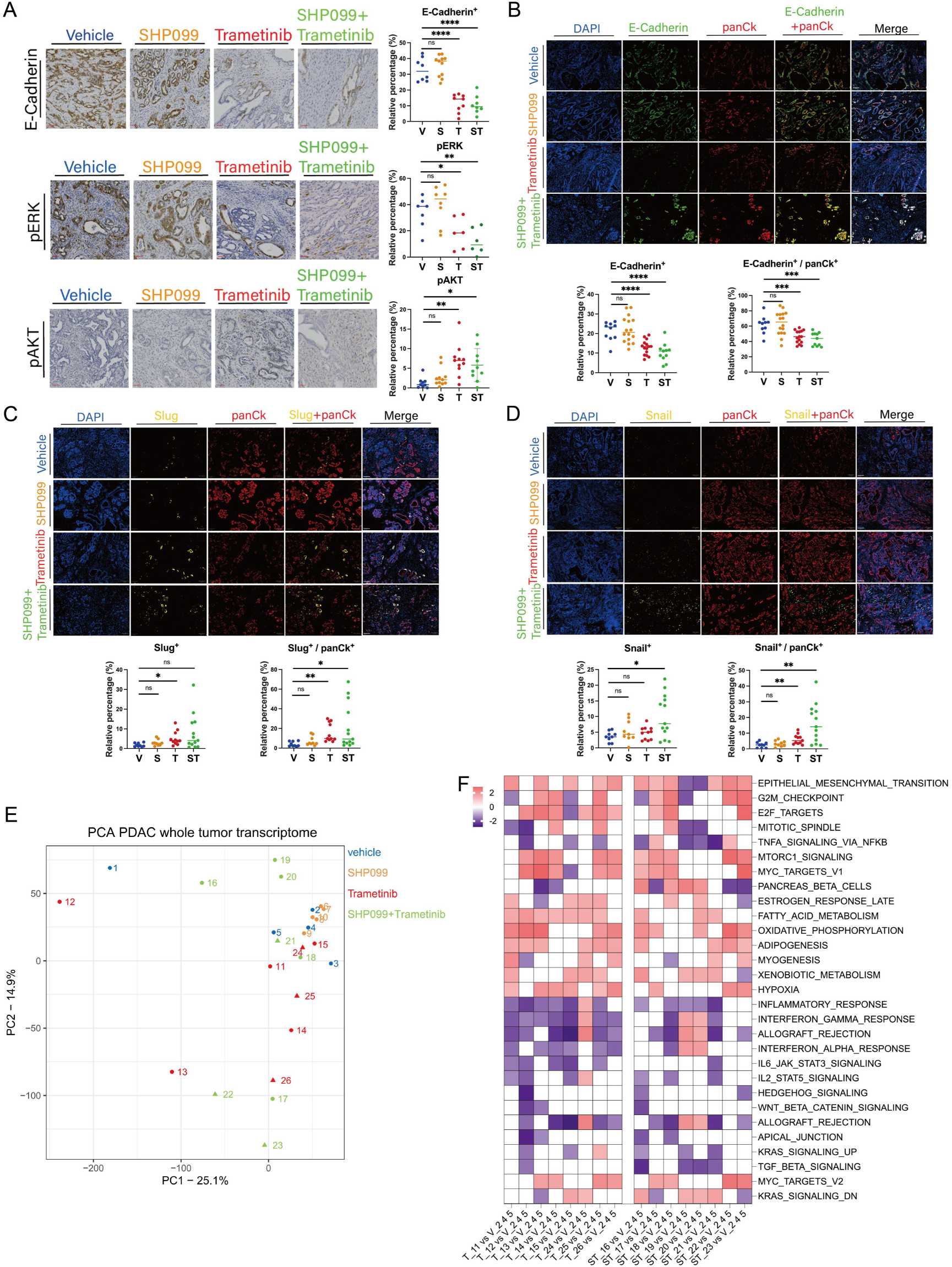
EMT accompanies evolving resistance to SHP2/MEK inhibition *in vivo*. (**A**) Conventional immunohistochemistry of pancreas tissues from mice of the different treatment arms of the survival cohorts with the antibodies indicated. Scale bars: 100 μm. (**B-D**) Multiplex immunohistofluorescence with pancreatic tissue of KPC mice from survival treatment cohorts. Scale bars: 100 μm. Representative images are shown. Statistical significance was determined via one-way ANOVA in panels B, C, D, with comparisons made against corresponding vehicle controls. **** p<0.0001; *** p<0.001; ** p<0.01; * p<0.05; ^ns^ p>0.05. (**B**) Tumor cell specific expression of E-Cadherin. V: n=10; S: n=16; T: n=15; ST: n=11. (**C**) Tumor cell specific expression of Slug. V: n=9; S: n=9; T: n=11; ST: n=13. (**D**) Tumor cell specific expression of Snail. V: n=9; S: n=9; T: n=11; ST: n=13. (E+F) Bulk tissue RNA Seq of tumor samples from mice of the different treatment arms of the survival cohorts. (E) PCA of KPC tumor transcriptomes following therapy. Vehicle samples 2, 4 and 5 cluster closely together, and near to all SHP099 samples, indicating the most prevalent “natural” endpoint of KPC tumors, qualifying them as a grouped reference. (**F**) Heatmap showing NES of MSigDB Hallmark gene sets for the comparisons of individual samples Trametinib (T) and SHP099+Trametinib (ST) to the grouped reference (merged vehicle samples 2, 4 and 5). Gene sets with adj. p < 0.05 are indicated.

To assess changes in tumor differentiation, we first analyzed H&E-stained pancreatic tumors from both short-term and survival endpoint cohorts. Tumors from vehicle– and SHP099-treated groups mostly retained well-differentiated, glandular architecture characteristic of low-grade (G1/G2) lesions. Remarkably, short-term treatment with Trametinib or dual SHP2/MEK inhibition appeared to further favor G1 histology, suggesting a partial reversal of early steps of dedifferentiation in the context of short-term therapeutic efficacy. In contrast, tumors exposed to prolonged treatment underwent a clear transition toward poorly differentiated, high-grade (G3) morphology, particularly in the Trametinib and dual combination groups (**Supplementary Figure 6A**, **B**).

To directly assess EMT activation during therapy response and resistance, we employed multiplex immunohistofluorescence. Tumor tissues were co-stained for E-Cadherin, the EMT transcription factors Snail or Slug, and pan-Cytokeratin (pan-CK) to ensure identification of dedifferentiated tumor cells that might otherwise resemble cancer associated fibroblasts. Whole-slide quantification in short– and long-term treated cohorts revealed that short-term dual SHP2/MEK inhibition led to a significant increase in E-Cadherin expression in pancreatic tissues, particularly within pan-CK⁺ tumor cells, confirming epithelial re-differentiation as a short-term response (**Supplementary Figure 7A**). At this stage, no significant changes in Snail or Slug expression were detected (**Supplementary Figures 7B**, **C**), suggesting that not necessarily mesenchymal-to-epithelial transition but reversal of early oncogenic processes may take place in the early phase of this model, where established cancer cells and multifocal tumors are still surrounded by premalignant lesions throughout the pancreas.

In contrast, long-term treated tumors exhibited significant E-Cadherin loss within pan-CK⁺ tumor cells from both MEKi and SHP2/MEKi cohorts compared to controls (**Figure 5B**). Concurrently, a significant upregulation in the expression levels of the EMT transcription factors Slug and Snail was observed in pan-CK^+^ tumor cells progressing under MEK and dual SHP2/MEK inhibition (**Figure 5C**, **D**). To validate these findings, we revisited our previously published KPC study using an earlier SHP2 inhibitor (GS493) in combination with Trametinib ^22^, and performed the same analysis with these historical samples. Notably, this additional survival cohort confirmed loss of E-Cadherin and increased expression of Snail and Slug, with more pronounced effects under dual inhibition (**Supplementary Figures 7D-F**).

Next, we performed RNA sequencing on bulk tumor tissues collected at the humane endpoint of the SHP099/Trametinib KPC trial. PCA of the transcriptomes showed that three of five vehicle-treated samples clustered closely together (samples 2, 4 and 5), which were thus set as a reference (**Figure 5E**, **Supplementary Figures 8A-C**). We performed an *in silico* deconvolution which demonstrated that the main contribution to the bulk tissue transcriptomes came from tumor cells (**Supplementary Figures 9A**). Comparing the quite heterogenous single treatment samples of T and dual S/T-treated tumors to this reference revealed enrichment of EMT-signatures as one of the most significant alterations in the majority of comparisons (**Figure 5F**).

Among the canonical signaling pathways analyzed, PI3K-AKT was most consistently enriched (in 3/8 T and 4/8 S/T samples). Additional pathway enrichments – including JAK-STAT, Wnt, Hippo, and Rap1 – were observed in one or two T or S/T samples, suggesting individualized compensatory signaling adaptations (**Supplementary Figure 9B**). KEGG pathway analysis also highlighted frequent alterations in drug metabolism and immune-related processes in ∼50% of the treated tumors, warranting further exploration (**Figure 5F**, **Supplementary Figure 9C**).

We then again applied the cell state gene modules from Barkley et al. and Franca et al. to the long-term *in vivo* bulk tissue RNA-seq datasets. Barkley’s pEMT, Glandular, and Mesenchymal states were simultaneously enriched in nearly all samples regardless of treatment; and no clear pattern emerged from Franca’s resistance continuum modules. These results likely reflect the complexity of tumor heterogeneity and the inherent noise in bulk RNA-seq datasets derived from multicellular tumor tissue, including abundant cancer-associated fibroblasts and immune cells, which are well-known and abundant components of the KPC PDAC microenvironment (**Supplementary Figure 10A-C**).

In summary, these findings confirm that histological progression, dedifferentiation, and EMT accompany the emergence of resistance to prolonged MEK or dual SHP2/MEK inhibition *in vivo*. While MAPK signaling remains largely suppressed, compensatory activation, most consistently via PI3K-AKT, facilitates adaptive resistance.

### Resistance development *in vivo* alters molecular subtype and cell state

To gain a more granular understanding of resistance development and the evolution of tumor cell states under therapeutic pressure *in vivo*, we performed single-cell RNA sequencing. KPC tumors from mice treated with vehicle, SHP099, Trametinib, or dual SHP099/Trametinib – both at short-term and long-term timepoints – were harvested, dissociated into single-cell suspensions, and sorted into CD45⁺ and CD45⁻ fractions for separate sequencing. Following quality control and integration of the single-cell datasets, dimensionality reduction and clustering identified 17 distinct cell populations. Cell type identities were assigned based on established marker genes curated from the literature (**Figure 6A**, **Supplementary Figure 11A**). To investigate the tumor-intrinsic changes, we isolated tumor/epithelial cell clusters from the full dataset and performed subclustering. This revealed 11 distinct tumor cell clusters, of which 6 exhibited predominantly epithelial and 5 showed mesenchymal expression patterns (**Figure 6B**). The vehicle control sample was dominated by epithelial clusters and short-term treatment shifted contributions of different epithelial clusters. In contrast, long-term treatment led to a pronounced enrichment of mesenchymal tumor cell clusters (**Figure 6C**, **D**; **Supplementary Figure 11B**).

**Figure 6.**
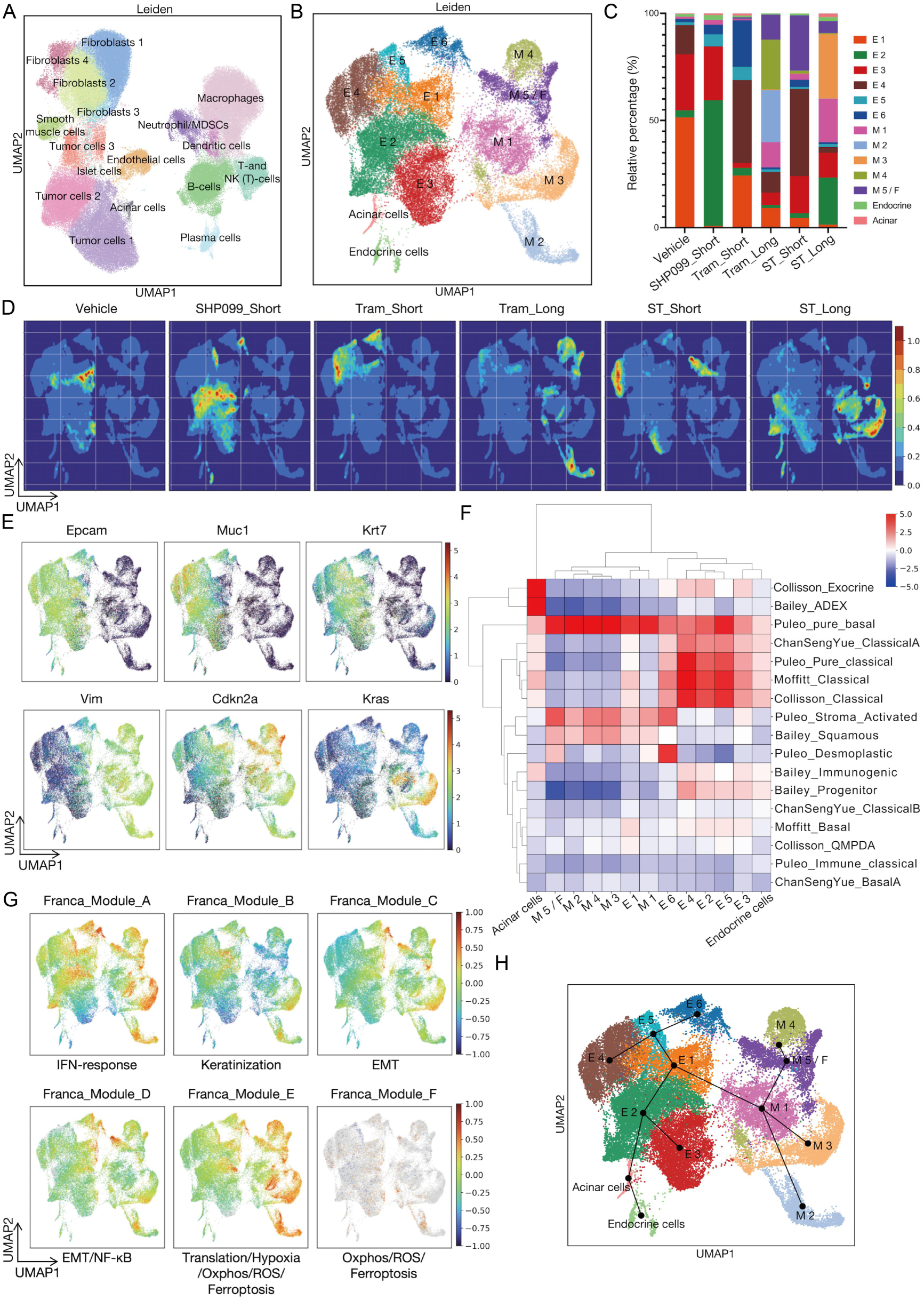
Single-cell transcriptomic profiling confirms EMT transition and PDAC subtype shifts under SHP2/MEK inhibition *in vivo*. (**A**) UMAP of all cells from murine tumors clustered by the Leiden algorithm, identifying 17 major populations including tumor cells, fibroblasts and immune cells. (**B**) Subset UMAP showing only tumor and epithelial cell clusters from A), reclustered for finer resolution. Thirteen tumor clusters were identified, including epithelial (E1–E6), mesenchymal (M1–M5/F), endocrine, and acinar cell clusters. (**C**) Stacked bar plot showing relative proportions of epithelial and mesenchymal cell clusters across treatment groups. “Short” indicating 2 weeks, “Long” indicating 5-6 weeks of *in vivo* treatment. Vehicle was administered for 2 weeks. (**D**) Density plots projecting treatment-specific cells onto the epithelial/mesenchymal UMAP space, revealing an expansion of mesenchymal cell states in the Trametinib and SHP099/Trametinib long-term treatment samples. (**E**) UMAP feature plots showing the expression of selected marker genes across all single cells. Feature plots display expression patterns of epithelial markers (*Epcam, Muc1, Krt7*), mesenchymal markers (*Vim, Cdkn2a*), and the oncogene (*Kras*). Color intensity represents log_2_-normalized expression levels, with higher expression shown in red and lower expression in blue. (**F**) Heatmap showing the enrichment scores of published PDAC molecular subtypes across the identified tumor/epithelial clusters (Acinar, Endocrine, E1–E6, M1–M5/F). Rows represent PDAC molecular subtype signatures from Collisson, Bailey, Moffitt, Puleo, and Chan-Seng-Yue classification systems. Color scale represents normalized enrichment scores (NES), with red indicating positive enrichment and blue indicating negative enrichment. (**G**) UMAP feature plots showing the distribution of Franca gene expression modules across all single cells: Module A (IFN-response), Module B (Keratinization), Module C (EMT), Module D (EMT/NF-κB), Module E (Translation/Hypoxia/Oxphos/ROS/Ferroptosis), and Module F (Oxphos/ROS/Ferroptosis). Scores are scaled per module, with red indicating higher and blue indicating lower scores. (H) Pseudotime trajectory visualization of the tumor cell subsets. Black lines indicate the predicted developmental paths between clusters based on pseudotime analysis.

Epithelial and mesenchymal tumor cell clusters were clearly separated by markers such as Epcam, Muc1, Krt7 and Vimentin (**Figure 6E**; **Supplementary Figure 11C**). Epithelial tumor cell clusters were significantly enriched for classical gene sets (Bailey “progenitor”, Chan-Seng-Yue “classical A”, Collisson “classical”, Moffitt “classical”, Puleo “pure classical”), while mesenchymal clusters were enriched for basal-like gene sets (Bailey “squamous”, Puleo “pure basal”, Puleo “stroma activated”, Puleo “desmoplastic”) (**Figure 6F**).

Applying an unsupervised clustering for enrichment scores for the cancer cell states from Barkley et al. ^28^ the epithelial clusters E1, E2, E4 and E5 grouped together, were associated with the Barkley glandular state, and overlapped with Bailey Progenitor and Puleo Pure Classical PDAC subtype enrichment scores. In contrast, the mesenchymal clusters M1-M3 grouped separately together, associated with the Barkley mesenchymal state and overlapped with Bailey Squamous and partly with Hwang Mesenchymal PDAC subtype enrichment scores. Of note, the partial EMT state signature, next to the stress module signature, was enriched in all tumor cell clusters (**Supplementary Figure 11D, E**).

We also again explored the gene modules associated with distinct transcriptional states of the resistance continuum described by Franca et al. ^29^. Enrichment for early resistance module A, dominated by IFN-response genes, was present in epithelial as well as mesenchymal clusters. The EMT-associated modules C and D, representing intermediate resistance states, were focally enriched in clusters and cells in or close to the mesenchymal space. Importantly, module E, representing the terminal resistance state S5 at the end of the described continuum, which is associated with metabolic adaptations including shifts toward oxidative phosphorylation, reactive oxygen species pathways and ferroptosis regulation, was strongly enriched in the mesenchymal clusters M1-M5, supporting the notion that M1-M5 represent late stage *in vivo* resistant tumor cells (**Figure 6G**).

We also performed a pseudotime trajectory analysis (**Figure 6H**), which revealed a dynamic progression originating from epithelial cluster E1, the dominant epithelial cluster in the DMSO vehicle sample (**Figure 6C**). One branch with direction towards E5 and then E4 or E6, and another with direction towards E2 and then E3 or the acinar/endocrine compartments, were associated with short term treatment, given the dominant contribution of the respective samples to those clusters (E5: SHP099 short, Trametinib short; E4: Trametinib short, SHP099+Trametinib short; E6: Trametinib short; E2: SHP099 short). Especially *Reg3b* and *Dmbt1* served as important contributors to the trajectory calculation (**Supplementary Figure 12A**) – their involvement in pancreatic epithelial regeneration and differentiation supporting short term treatment responses and re-epithelialization. A different trajectory originating from E1 crossed over to the mesenchymal side, with the EMT genes *Vimentin* and *Lgals1* as important contributors (**Supplementary Figure 12A**), first towards M1, and from there to M5 and M4, or M3 or M2. M1, as this common origin of long-term treatment resistance, was accordingly represented well in Trametinib long as well as SHP099+Trametinib long samples, while M2 and M4 were virtually specific to Tram long and M3 specific to SHP099+Trametinib long, respectively (**Figure 6B and Supplementary Figure 11B**). These trajectories underscored the differences in tumor cell plasticity in short term vs. long term SHP2/MEK-inhibition in vivo, with long-term treatment driving the emergence of resistant mesenchymal phenotypes. It is however also limited by the fact that it falls short to provide a trajectory-connection between the early and late response clusters (e.g. from E4 to M1).

Pathway enrichment analysis across BIOCARTA, KEGG, REACTOME, and Oncogenic Signature (MSigDB C6) gene sets confirmed gene sets related to PI3K-AKT, Rap1, TGFβ, Wnt/β-catenin and Hippo pathways enriched in mesenchymal tumor cell clusters. In the BIOCARTA collection, TGFB_PATHWAY was significantly enriched in mesenchymal cluster M3, while INTEGRIN_PATHWAY (upstream of PI3K-AKT and Rap1 signaling) was enriched in M3 and M4 (**Supplementary Figure 12B**). With the Oncogenic Signature (MSigDB C6) collection, Wnt/β-catenin transcription factor target signatures (LEF1_UP.V1_DN and LEF1_UP.V1_UP) were enriched in M1 and M4 and M5/F, while AKT-related gene sets were enriched in M1, M2 and M5/F (AKT_UP.V1_UP and AKT_UP.V1_DN) (**Supplementary Figure 12C**). REACTOME analysis further highlighted Integrin cell surface interactions and Cell-cell communication (Rap1/PI3K-AKT-related) in M3 (**Supplementary Figure 12D**). The KEGG analysis identified enrichment of ECM–receptor interaction and Focal adhesion (both closely linked to PI3K-AKT and Rap1) in M1 and M3 (**Supplementary Figure 12E**). In addition, mesenchymal clusters M1 and M3, representing roughly 50% of the tumor cells in the SHP099+Trametinib long samples, demonstrated upregulation of *Kras* expression (**Figure 6E**).

Overall, single cell RNA-sequencing findings confirmed that EMT and dedifferentiation accompany emergence of resistance and underscored molecular subtype shift and cell state transitions as hallmarks of the adaptive response of murine KPC PDAC to dual SHP2/MEK inhibition. They further supported a contribution of increased RAS expression and/or PI3K-AKT, Rap1, TGFβ, and partially Wnt/β-catenin signaling activation in adaptive resistance.

### Therapy response and patterns of resistance in patient-derived samples

For translational confirmation, we employed human PDAC cell lines and patient-derived PDAC organoids (PDO), representative of the whole spectrum from classical to basal-like subtypes, to establish and analyze human in *vitro* models of adaptive resistance to vertical RAS pathway inhibition.

Three well established PDAC cell lines (MIAPaCa II, PANC1, YAPC) were selected, based on their molecular subtype classification, validated through phenotypic and transcriptomic features, by assessment of morphology, subtype-associated protein expression and subtype-specific gene signature enrichment (**Figure 7A**, **Supplementary Figure 13A**). The cell lines were treated for 72h for short-term response assessment, and adaptive resistant lines were established by continuous treatment for at least 6 weeks.

**Figure 7.**
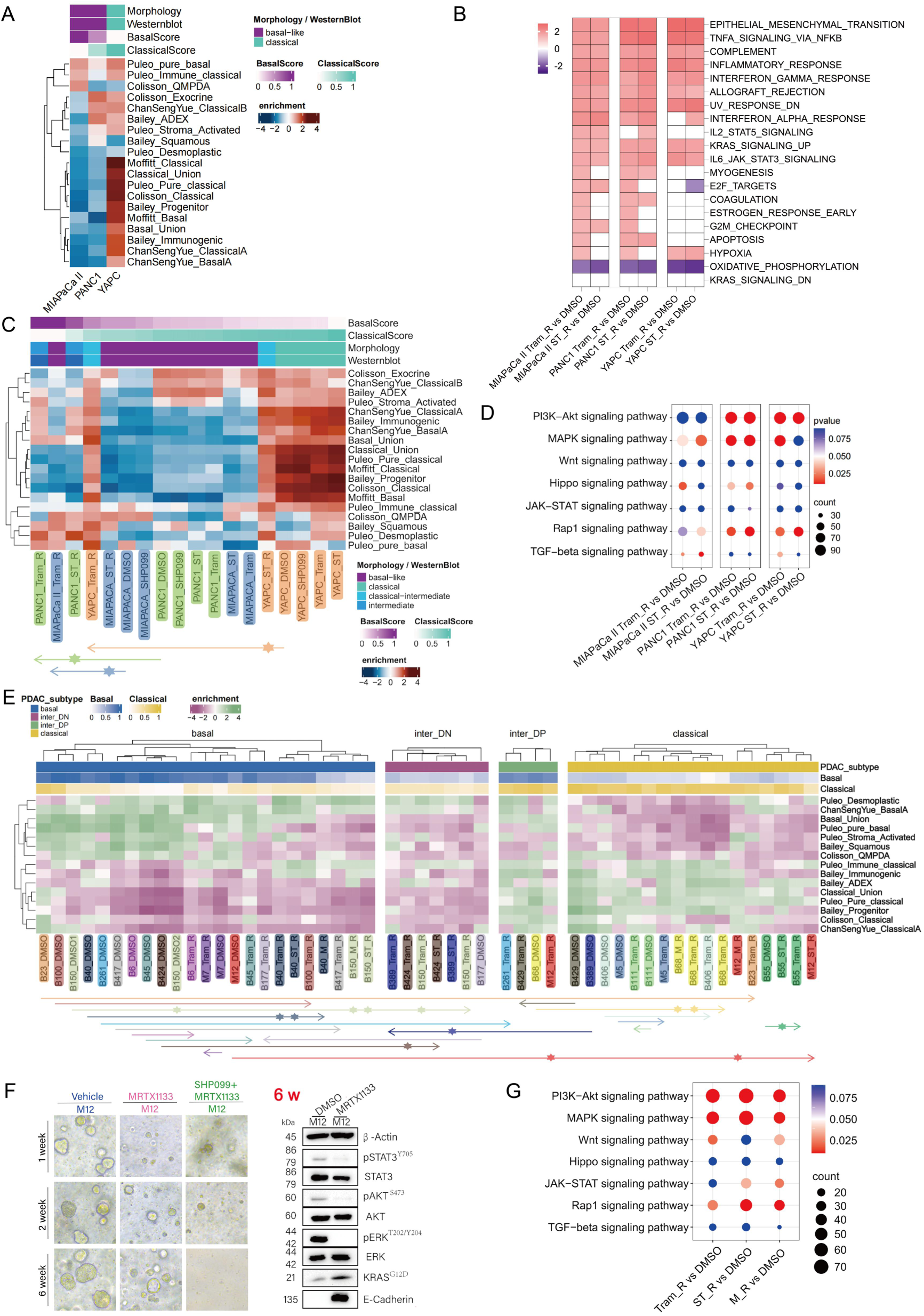
Therapy response and patterns of resistance in human cell lines and patient-derived PDAC organoids. (**A-D**) **Human PDAC cell lines**: (**A**) Heatmap showing enrichment scores for published PDAC subtype signatures across human-derived pancreatic cancer cell lines, annotated with morphological classification (basal-like, classical, intermediate), Western blot markers, and calculated Basal and Classical subtype scores. (**B**) Hallmark gene set enrichment analysis comparing resistant versus vehicle-treated cells. Gene sets with adj. p < 0.05 are indicated. (**C**) Heatmap of subtype signature scores across resistant and sensitive human cell lines treated with various inhibitors. Each arrow connects different treatment conditions of the same cell line. Arrows with identical colors indicate subtype transitions within the same cell line across treatments, reflecting therapy-induced transcriptional reprogramming. (**D**) KEGG GSEA dot plot illustrating pathway enrichment across cell lines with resistance to MEK inhibition or dual SHP2/MEK inhibition. Dot size indicates gene count; color reflects statistical significance (adjusted p-value). (**E-G**) **Patient derived PDAC organoids** (**PDO**): (**E**) Subtype-resolved enrichment heatmap for Basal-like (Basal), Intermediate double-negative for Basal and Classical (InterDN), Intermediate double-positive for Basal and Classical (InterDP), and Classical subtypes. The heatmap included treatment resistant samples, illustrating transcriptomic shifts in resistant conditions. Each arrow connects different treatment conditions of the same organoid line. Arrows with identical colors indicate subtype transitions within the same PDO line across treatments, reflecting therapy-induced transcriptional reprogramming. (**F**) Light microscopic appearance and immunoblot assays with the indicated antibodies PDAC patient-derived organoid M12 treated long-term with KRAS^G12D^ and SHP2/KRAS^G12D^ inhibition for 6 weeks; no viable cells and consequently no protein sample was recoverable from the dual SHP2/KRAS^G12D^ inhibition approach). Scale bars: 200 μm. β-Actin served as the loading control. (**G**) Key signaling pathway analysis in resistant PDAC patient-derived organoids. Transcriptome data from individual samples were merged according to their treatment (DMSO: n=19, Tram_R: n=19, ST_R: n=19, M_R: n=4). Dot size indicates gene count; color reflects statistical significance (adjusted p-value).

In YAPC cells, which display an epithelial morphology and represent a more classical subtype, pronounced features of epithelial–mesenchymal transition (EMT) were observed and confirmed in response to prolonged MEK inhibition and SHP2/MEK inhibition. These included a morphological shift toward a mesenchymal phenotype, marked downregulation of the epithelial marker E-cadherin, and complete loss of GATA6 expression, a transcription factor strongly associated with the classical PDAC subtype. Despite the development of treatment resistance, ERK phosphorylation remained suppressed. In contrast, RAS expression, as well as AKT and STAT3 activation, were upregulated, with more pronounced effects observed under MEK inhibition compared to dual SHP2/MEK blockade (**Supplementary Figure 13B**).

PANC1, a cell line with mixed epithelial and mesenchymal traits and signature enrichment scores placing it in the intermediate range of subtypes, the morphological phenotype remained largely stable following treatment. However, resistance to MEK or dual SHP2/MEK inhibition was accompanied by upregulation of E-Cadherin, and downregulation of Vimentin and Slug, suggesting partial epithelial re-differentiation. ERK phosphorylation again remained suppressed, with the most sustained inhibition achieved under dual therapy. STAT3 phosphorylation and β-Catenin expression were increased with both MEK and dual inhibition resistance, indicating a potential compensatory survival mechanism (**Supplementary Figure 13C**).

MIAPaCa-II cells, representing a basal-like, mesenchymal subtype, retained their mesenchymal phenotype following MEK or SHP2/MEK inhibition. E-cadherin expression remained completely absent, while N-cadherin and Vimentin levels showed only slight reductions. Notably, MIAPaCa-II cells demonstrated the ability to reactivate ERK signaling under resistant conditions, whereas STAT3 activation appeared to contribute only to early adaptive responses (**Supplementary Figure 13D**).

In PANC1, the only cell line of these three with a KRAS^G12D^ mutation, we further investigated the effects of KRAS^G12D^ inhibition. In a similar fashion microscopic phenotype was retained, a marked upregulation of E-Cadherin, especially with SHP2/ KRAS^G12D^ resistance was accompanied by retained Vimentin levels. ERK phosphorylation, AKT phosphorylation and compensatory RAS-expression was best suppressed with the combination therapy, with upregulation of β-Catenin again as potential major survival signal (**Supplementary Figure 13E**).

To further investigate transcriptional changes associated with treatment resistance, RNA sequencing was performed on all three PDAC cell lines across treatment conditions. Hallmark gene set collection analysis revealed consistent and significant enrichment of EMT signatures in all resistant cell populations (**Figure 7B**). Single-sample GSEA using subtype-specific gene sets showed that MEK or dual SHP2/MEK inhibition consistently induced a shift towards the basal-like end of the subtype spectrum, particularly and most prominently in YAPC (**Figure 7C**). Cell state analyses based on the Barkley et al. and Franca et al. gene modules demonstrated pEMT as the dominant pattern, already without treatment, and an increased enrichment of Cycle and Basal cell states in dual inhibition resistant samples (**Supplementary Figures 14A**, **B**, **C**). Pathway gene set enrichment analysis in part confirmed results from protein analyses above and highlighted cell line-specific patterns of compensatory pathway activation during resistance development: PI3K-AKT and Rap1 in YAPC; PI3K-AKT, Rap1, MAPK and Hippo in PANC1; and MAPK and TGF in MiaPaCa-II (**Figure 7D**).

We then utilized over 40 PDO lines representing the spectrum of classical to basal-like transcriptional subtypes. These PDOs were treated over extended periods of time and multiple passages with either T, S/T, MRTX1133 (M) or SHP099 + MRTX1133 (S/M) until they resumed stable proliferation.

In general, treatment efficacy of (vertical) RAS pathway inhibition was high across these 40 PDO lines, and we thus were able to successfully establish treatment resistance in less than 50%, i.e., only 19 of these 40 PDO lines (all 19/19 resistant to Trametinib: T_R; 7/19 resistant to S/T: ST_R; 4/19 resistant to M: M_R; and none (0/19) resistant to S/M; **Supplementary Figure 15A-F**). The resistant lines, along with their corresponding parental lines (untreated (UT) and/or treated with DMSO vehicle control) were subsequently subjected to RNA sequencing.

Single-sample GSEA for transcriptional PDAC subtyping yielded a spectrum ranging from basal-like through intermediate (double-negative/low: DN, or double-positive/low: DP, for classical and basal-like ssGSEA) to fully classical subtypes (**Figure 7E**). To our surprise, among the 19 resistant PDO lines, only 3 shifted toward the basal-like end of the spectrum relative to their respective parental lines: two from classical to intermediate (B389, B429) and one from intermediate to basal-like (B177). In contrast, six lines exhibited an opposite shift toward the classical phenotype: two from basal-like to classical (B023, M12), three from basal-like to intermediate (B150, B261, B424), and one from intermediate to classical (B068). The remaining 10 lines, i.e. the majority, retained their original subtype classification, comprising six basal-like (B006, B40, B45, B100, B417, M7) and four classical (B55, B111, B406, M5). Overall, the mean transcriptional trajectory pointed rightward on the heatmap, indicating a general shift toward a more classical phenotype, particularly among originally basal-like lines (**Figure 7E**). These observations were corroborated in selected PDO lines, such as M12 and B150: M12 organoids resistant to MRTX1133, T or ST displayed more abundant spherical, hollow morphology compared to the compact, lumen-less “snowball” architecture of the parental M12 line (**Supplementary Figure 15B**). This was accompanied by a marked upregulation of E-Cadherin. In parallel, ERK phosphorylation was efficiently suppressed (**Figure 7F**, **Supplementary Figure 15C**). In contrast, B150 organoids resistant to MRTX1133 exhibited a loss of E-cadherin expression (**Supplementary Figure 15D**). Interestingly however, B150 resistant to T or ST had a phenotype comparable to resistant M12 lines, with a shift from rather solid to mixed shape and strong increase in E-Cadherin levels, accompanied by pERK suppression and compensatory AKT activation (**Supplementary Figure 15E**). These findings were further supported by immunofluorescence analyses, which confirmed treatment-induced changes in EMT-protein expression (**Supplementary Figure 15G**). Notably, SHP2/KRAS^G12D^ co-inhibition completely abrogated the outgrowth of viable resistant organoids across the whole panel, underscoring its superior therapeutic efficacy. Transcriptomic GSEA pathway analysis comparing groups of T, ST and MRTX1133 resistant lines with their DMSO treated controls again revealed PI3K–Akt, MAPK, Wnt, JAK–STAT, and Rap1 signaling as potential mediators of adaptive resistance (**Figure 7G**).

Obviously, the pattern of adaptive reprogramming observed across a broad range of PDAC PDOs contrasted to some extent with the responses seen in 2D *in vitro* systems and our KPC *in vivo* model, suggesting a more complex and heterogenous landscape. However, the specific culture conditions used for PDOs, including tailored media formulations and the Matrigel matrix, may preferentially support epithelial, differentiated cell states, or clones, i.e., classical-like subtypes ^31,32^. Consequently, the observed transcriptional shifts may not exclusively reflect intrinsic subtype plasticity, but could also result from clonal selection from a heterogeneous population under therapeutic pressure, within a microenvironment that favors epithelial identity, thus limiting the interpretability of PDO findings.

In summary, these experiments with human PDAC models confirmed the enhanced efficacy and translational potential of SHP2 inhibitor-based combination therapies, both in conjunction with MEK and particularly KRAS^G12D^ inhibition. The data also underscored the propensity of vertical RAS pathway targeting to induce cell state transitions. While 2D human culture models largely supported the general concept of dedifferentiation and a shift toward basal-like subtype, our 3D PDO models suggested that more heterogeneous response patterns might have to be anticipated. Yet, clonal selection within the specific PDO culture setup may have introduced bias and limited generalizability. Finally, the findings emphasize that compensatory signaling during adaptive resistance is highly context-dependent and subject to intertumoral variability.

## Discussion

RAS pathway inhibition holds renewed promise for transforming the clinical management of patients with pancreatic cancer. The development of direct RAS inhibitors represents a major breakthrough, given the critical dependence of PDAC on this central and highly prevalent oncogene, the downstream signaling cascades it activates, and the pivotal role of the ERK-dependent transcriptome ^33^. However, both intrinsic and adaptive resistance limit the efficacy and durability of these agents. Vertical pathway inhibition leveraging the properties of the tyrosine phosphatase SHP2, which integrates signals from virtually all receptor tyrosine kinases to the RAS-RAF-MEK-ERK signaling cascade ^26^, has emerged as a promising strategy to overcome adaptive resistance to MEK or ERK inhibition ^22,25^. For the same reasons, SHP2 inhibition may also enhance the effectiveness of direct RAS-targeted therapies ^24^.

Building on observations of adaptive resistance to dual SHP2 and MEK inhibition in a murine genetically engineered murine PDAC model, this study provides insights into phenotypes and mechanisms of resistance to SHP2-based vertical RAS-pathway inhibition, stratified by PDAC transcriptional subtypes. Our findings reveal shifts in differentiation/EMT status, transcriptional subtype and tumor cell states as a common pattern of resistance across both murine and human PDAC models. These adaptations are accompanied by heterogeneous alterations in compensatory signaling pathways, underscoring the need for individualized precision medicine combination partners or, more general, strategies exploiting resistant cell-state inherent vulnerabilities, in future clinical trial design.

Primary resistance to SHP2-based vertical RAS inhibition was virtually absent, with potent (initial, short-term) therapeutic responses observed across the full spectrum of PDAC transcriptional subtypes and across species. Notably, neither transcriptional subtype or differentiation state per se did predict treatment response. Interestingly, treatment-naïve basal-like cells exhibited marked sensitivity, challenging the long-held assumption that epithelial PDAC cells exhibit greater RAS dependency. The cell state emerging with adaptive resistance *in vitro* and *in vivo* in most instances featured basal-like transcriptional subtype shift, dedifferentiation, mesenchymal shape, and EMT expression profile. Associated with this process, the context-dependent activation of several compensatory signaling pathways, including RAS-RAF-MEK-ERK, PI3K-AKT, JAK-STAT, Wnt/β-catenin, and Rap1, was observed. Importantly, dual SHP2 and KRAS^G12D^ (MRTX1133) inhibition showed strong therapeutic potential, since generating resistant cell lines or patient-derived organoids to this combination proved notably difficult.

Together, these data reiterate a number of the previously observed and reported concepts in the realm of adaptive treatment resistance but also add important insight into SHP2-based vertical RAS-pathway inhibition approaches. First, EMT, differentiation shifts and cell plasticity have been described as phenomena accompanying treatment resistance to chemotherapy and targeted therapies in general, ^32,34–37^ and KRAS^G12C^^38,39^ and KRAS^G12D^ inhibition ^21^ in particular. In more detail, EMT induction was ascribed an important contribution to partial resistance and thereby, more generally, facilitating phenotypic plasticity over the long term, enabling the emergence of additional mechanisms that increase drug resistance along a resistance continuum ^29^.

RAS-amplification and increased RAS expression ^27^, compensatory activity of YAP/TEAD ^40^ and Pi3K/AKT/mTOR-signaling ^21,27,41^ have been implicated in the context of direct RAS inhibition as a single agent. In a study investigating resistance of non-small-cell lung cancer to combined SHP2 and KRAS^G12C^ inhibition, similarly, KRAS^G12C^ amplification and an exploitable dependency on Pi3K signaling was observed ^27^.

There are several limitations in this study. First, information regarding genetic and epigenetic alterations with adaptive resistance in our models and treatment combinations is lacking, and mechanistic insights are therefore non-comprehensive. Second, we here only utilized a mutant-specific OFF-state KRAS inhibitor and (newer) compounds mutant-selectively targeting GTPase ON-states and/or non-mutant-selectively targeting all KRAS or RAS molecules or isoforms, respectively, may even be superior ^6,42^ in potency. We however anticipate similar resistance patterns to evolve. Third, we here did not combine RAS-pathway inhibition with standard-of care chemotherapy, falling short of providing insight into the potential of the most obvious and most simple combinatorial clinical application. Finally, organoid culture technology with its proclivity for epithelial/classical cell states ^31^ and its variability due to the dependency on the specifics of the matrix components used ^43^, impairs generalizability of experimental results.

Nonetheless, or work confirms and extends central aspects of the properties and mechanisms of adaptive resistance to RAS-MAPK pathway inhibition. Our discoveries highlight SHP2 as a compelling combination partner not only for MEK but also for direct RAS inhibitors, potently delaying the onset of resistance. Moreover, our data provide a foundation for future efforts to enhance the durability of vertical RAS-pathway inhibition and sustainably improve patient outcomes. Exploiting context-dependent compensatory signaling alterations in response to vertical RAS-inhibition will require personalized medicine approaches and bears the risk of another round of swift re-adaptation. Strategies aimed at resistant mesenchymal cell states such as ferroptosis induction ^44,45^ or impairment of the unfolded protein response ^46^ may be of more general value. Finally, more effort must be devoted to better understanding and harnessing the cross-talk between resistant cell populations and the immune response – likely the only agent capable of achieving long-term control, or even a cure.

## Materials and Methods

### Mouse model

Kras^tm1Tyj^: *Kras^LSL-G12D/+^* + Trp53^tm1Brn^: *Trp53^fl/fl^* + Ptf1a^tm1(Cre)Hnak^: *Ptf1a^Cre-ex^*^1^*^/+^* (KPC) compound mutant mice with a mixed genetic background were used. Mice were kept in a specific pathogen-free facility. KPC mice develop a spectrum of premalignant lesions called Pancreatic Intraepithelial Neoplasia (PanINs) that ultimately progresses to overt and multifocal ductal adenocarcinoma with 100% penetrance. The tumors are heterogenous, but generally have a moderately differentiated ductal morphology from the outset, with extensive stromal desmoplasia. (Parts of the) tumors may progress to higher-grade, dedifferentiated lesions over time. Genotypes were determined by PCR and gel electrophoresis at weaning and after death. All experiments were conducted at the facilities of the Center for Experimental Models and Transgenic Services (CEMT) at the Medical Center University of Freiburg, Germany. The animals were maintained in hygienic, pathogen-free conditions, and all animal experiments were approved by the local authority (Regierungspräsidium Freiburg, Germany; approval number G/19-137). The authors complied with the “Animal Research: Reporting of *In Vivo* Experiments” (ARRIVE) guidelines.

### Drugs and inhibitors

The drugs SHP099 (S8278) and Trametinib (S2673) were purchased from Selleckchem. MRTX1133 (HY-134813) was obtained from Medchemexpress LLC. Drugs were dissolved in DMSO to yield 5-50mM stock solutions and stored at –80°C.

### In *vivo* therapy dosing

KPC mice were screened for established PDAC tumors by palpation and weekly MRI. Upon tumor detection, tumor-bearing mice were randomly divided into four cohorts and received vehicle, SHP099 (75 mg/kg, p.o., qod), Trametinib (1 mg/kg, p.o., qod) or combination (SHP099, 75 mg/kg, p.o., qod; Trametinib, 1 mg/kg, p.o., qod) for the indicated period of time. The vehicle was composed of 4% DMSO, 0.2% Tween-80, and 5% glucose in ddH₂O. All formulations were freshly prepared and kept cool prior to use and administered by oral gavage at a dosing volume of 5 mL/kg body weight. The tumor volume was monitored by MRI scanning every week.

### MRI

The KPC mice mentioned above were subjected to MRI scanning experiments once a week. Sedation was performed via continuous inhalation of 2% isoflurane (Abbott) in O2 using a veterinary anesthesia system (Vetland Medical). Body temperature was maintained and monitored, and eyes were protected by eye ointment. Scans were performed on a Bruker 9.4-T BioSpec system (BrukerCorp., Ettlingen, Germany) using two separate mouse-volumequadrature-resonators (Bruker Corp., Ettlingen, Germany) with identical geometry (inner diameter 38 mm). Solid tumor volumes were calculated using the medical imaging platform NORA by summating truncated pyramid volumes between tumor areas on vicinal slices.

### Antibodies

Proteins were detected by western blot with the following antibodies: pSTAT3 1:1000 (D3A7), STAT3 1:1000 (79D7), SHP2 1:1000 (D50F2), pAKT^S473^ 1:1000 (D9E), AKT 1:1000 (C67E7), pERK^T202/Y204^ 1:1000 (D13.14.E4), ERK 1:1000 (137F5), KRAS 1:1000 (E2M9G), KRAS^G12D^ 1:1000 (D8H7), ZEB1 1:1000 (D80D3), E-Cadherin 1:1000 (24E10), N-Cadherin 1:1000 (D4R1H), Vimentin 1:1000 (D21H3), GATA-6 1:1000 (D61E4), Slug 1:1000 (C19G7), Snail 1:1000 (C15D3), β-Catenin 1:1000 (D10A8), KRT81 1:1000 (H00003887-M01J) and Actin 1:5000 (A2066). All antibodies were purchased from Cell Signaling Technologies except KRT81 (Thermo Fisher Scientific) and Actin (Sigma-Aldrich).

### Histology

Tissue specimens were fixed in 4% buffered paraformaldehyde, dehydrated and embedded in paraffin wax. Formalin-fixed paraffin-embedded sections of 3µm were stained with H&E, Sirius Red or used for immunohistochemical studies.

Immunohistochemistry was performed on murine formalin fixed paraffin-embedded sections using avidin–biotin enhancement (Vector Laboratories). The following antibodies were used: pERK1/2 (4370; 1:1000), pAKT (4060; 1:100), E-Cadherin (3195; 1:800), Slug (9585; 1:100), Snail (3879; 1:100), Ki67 (9449; 1:800) from Cell Signaling, pan-Cytoleratin (ab7753; 1:500) from Abcam, Berlin, Germany. IHC slides were developed with DAB (Liquid DAB+ Substrate Chromogen System, Dako) and counterstained with hematoxylin. For immunofluorescence (IF), DAPI was included for identification of nuclei and pan-cytokeratin for identification of epithelial cells. Secondary Opal polymer HRP anti-mouse and anti-rabbit antibodies and Opal fluorophores were used to detect primary antibodies. Primary antibodies were first optimized via immunohistochemistry on control tissue to confirm contextual specificity. Monoplex immunofluorescence and iterative multiplex fluorescent staining were then used to optimize staining order, antibody-fluorophore assignments, and fluorophore concentrations.

Image acquisition was achieved on a Zeiss AxioImager or the-PhenoCycler-Fusion 2.0 Solution. Quantitative analyzes of tumor areas and immunohistochemistry staining were performed with Qupath and ImageJ softwares.

### Cell culture and cell lines

Primary murine PDAC cells were derived from tumors of freshly sacrificed KPC mice. The tumors were prepared as quickly as possible, and 3-4 tumor pieces of approximately 5mm were cut into small fragments using a scalpel. These tissue pieces were cultured in RPMI 1640 supplemented with 10% FCS and 1000 U/ml Penicillin-Streptomycin. After detecting cell growth in a two-dimensional layer, any remaining tissue remnants were removed by trypsinization and the cells were recultured.

The human PDAC cell lines MIA PaCa II, PANC-1, and YAPC were obtained in-house and cultured in RPMI 1640 supplemented with 10% FCS and 1000 U/ml Penicillin-Streptomycin.

All cells were handled under sterile conditions and kept at 37 °C in a humidified incubator with 5% CO_2_.

### Immunoblotting

Cells were lysed with RIPA buffer and protein concentrations determined. Lysates were denatured, loaded onto SDS-PAGE, and transferred to nitrocellulose. After blocking, membranes were incubated with primary and secondary HRP-conjugated antibodies, then visualized with ChemiDoc.

### RAS-RAF-RBD pulldown

Cells were lysed in Mg^2+^ lysis buffer (125 mM HEPES, pH 7.5, 750 mM NaCl, 5% Igepal CA-630, 50 mM MgCl2, 5 mM EDTA and 10% glycerol; Millipore) supplemented with protease inhibitor and phosphatase inhibitor cocktails. RAS-GTP levels were measured using the RAS Activation Assay Kit from Millipore (17-218) per manufacturer’s instructions. Briefly, fresh pancreatic tissue or PDAC cell lines were lysed in ice-cold Mg2+ lysis buffer and equal amounts of protein were incubated with RAF –1-RBD agarose beads for 45 min at 4 °C on a rotator. After three washing steps, beads were suspended in Laemmli-reducing sample buffer, subjected to SDS–PAGE and blotted on nitrocellulose membranes. Detection was performed with the indicated antibodies.

### Generation of drug-resistant cell lines

In order to obtain cell lines resistant to four different conditions, cell lines were treated with DMSO, SHP099, Trametinib, MRTX1133, and the combinations SHP099+Trametinib and SHP099+MRTX1133, with fixed drug concentrations until they resumed steady proliferation, i.e. for 6-10 weeks. Samples were collected at 90% confluence as described above. The obtained samples were then used for RNA sequencing and immunoblotting assays.

### Transcriptomics

To investigate the transcriptomic behavior of human and murine PDAC 2D cells the cells were cultured with a corresponding density that reaches 90% at the end of experiment for each treatment arm. The cells were plated and cultured in RPMI 1640 with 10% fetal calf serum (FCS) and 1000 U/ml penicillin-streptomycin overnight to allow attachment to the surface area. After 12-24 hours, we added either 10mM of SHP099, 20nM of trametinib, and a combination of both or 10uM MRTX1133, and a combination of SHP099 and MRTX1133 as targeted therapy, in triplicate each. After 48 hours the cells were lysed in RLT Plus buffer from QIAGEN and subsequently purified via RNeasy® Plus Mini Kit (QIAGEN 74136). PDO cells were recovered from Matrigel domes and processed in analogy. PDOs were collected in untreated or DMSO control treatment states, and following a prolonged period of treatment for at least 6 weeks over multiple passages. KPC tissue samples were collected from survival mice and lysed and purified as above.

Sequencing was conducted at Deutsches Krebsforschungszentrum Heidelberg (DKFZ) using Illumina’s NovaSeqTM6000. Data were converted to FASTQ files, quality-trimmed, and aligned to reference genomes (GRCh38 for human, GRCm39 for mouse) with STAR aligner (v2.7.10a) ^47,48^. Triplicate sample data for 2D cell line each condition were merged. PDO samples were individual samples.

Differential expression analysis was performed using limma package, followed by gene set enrichment analysis (GSEA) with the fgsea algorithm and gene-sets from the MSigDB collection ^49–51^. Differentially expressed genes (DEGs) were identified from RNA-sequencing datasets using the thresholds of adjusted p-value < 0.05 and absolute log₂ fold change >= 1, and subsequently subjected to pathway analysis. Kyoto Encyclopedia of Genes and Genomes (KEGG) pathways were analyzed employing the clusterProfiler R package ^52^, while pathway-based visualization was performed using the pathview package ^53^. Multiple testing correction was applied using the Benjamini–Hochberg method to control the false discovery rate (FDR). PDAC subtypes were classified based on five key studies (Moffitt, Bailey, Collisson, Puleo, Chan-Seng-Yue), combining subtype signatures into Basal and Classical categories. To facilitate integrative analyses, we defined two overarching categories – basal union and classical union – by consolidating subtype signatures corresponding to the basal-like and classical phenotypes, respectively. Subtype-specific scores were computed for each (untreated) sample via single-sample gene set enrichment analysis (ssGSEA), implemented through the clusterProfiler package ^54^. Prior to ssGSEA, gene expression values were scaled across the entire cohort. Human PDAC signatures were mapped to their murine orthologs via the homologene R package.

### Single-cell sequencing

Treated KPC tumors were harvested, minced, digested with collagenase II (C6885, Sigma, 5 mg/mL in RPMI) for 20 min at 37°C, and filtered through a 100 µm and 70 µm cell strainer. The cells were stained with anti-CD45 AF700 (BioLegend, 103128) for 20 min on ice and sorted via the BC Cytoflex SRT. For single-cell mRNA barcode labeling, the GEXSCOPE® Single-cell RNA Library Kit Cell V2 Kit from Singleron Biotechnologies GmbH was used, and cDNA was sent for library preparation and sequencing via an Illumina PE 150. Alignment and quantification of single-cell RNA-seq data were performed using CeleScope (Singleron Biotechnologies, version 1.14.0) and aligned to the mouse genome “Mus_musculus_ensembl_92” (Ensembl). Count matrices were analyzed with Scanpy (version 1.9.1) ^55^. Quality control excluded cells with fewer than 500 genes, more than 25,000 UMI counts, or over 10% mitochondrial counts. Gene expression was log-normalized and scaled by 10,000 ^56^. PCA was performed on the top 5,000 variable genes ^57^. Leiden clustering (resolution 0.5) was used, and UMAP was employed for visualization ^58,59^. Differentially expressed genes (DEGs) were identified using Wilcoxon rank-sum test with adjusted p-values <= 0.05 and log fold change >= 1. GO enrichment analysis was performed with ClusterProfiler in R ^52,54,60^ (version 4.2.2) for curated gene sets ^28^ in scRNA-seq data, which were visualized in UMAP.

### Patient-derived ex vivo PDAC organoids

Human PDAC tissues were obtained from surgical resections performed at the Department of General and Visceral Surgery, Center for Surgery, Medical Center – University of Freiburg, Faculty of Medicine, Freiburg, Germany. All patients provided written informed consent prior to surgery, and tissue collection was in accordance with the Declaration of Helsinki and approved by the institutional Ethics Committee of the University of Freiburg (No. 126/17 and No. 73/18). PDAC diagnosis was confirmed by routine pathology after resection of pancreatic tumors. Fresh PDAC tissues were minced, collagenase-treated, and ACK-lysed to isolate cells, which were then cultured as organoids in Matrigel domes with human feeding medium, following protocols adapted from the Tuveson Lab ^61^.

### Statistical analysis

Kaplan–Meier survival curves were calculated from all individual survival times of mice from the different cohorts. Curves were compared by log-rank (Mantel–Cox) test to detect significant differences between the groups. For image quantifications, significance was assayed by unpaired, two-tailed Student’s t-test or Mann–Whitney test for comparison of two groups and by one-way analysis of variance (ANOVA) with post-hoc Tukey’s test for more than two groups; ****P<0.0001; ***P<0.001; ** P<0.01; *P<0.05; ^ns^ p>0.05. Statistical analysis was performed with GraphPad PRISM 9.4.1 software.

## Acknowledgements

The authors thank the Core Facility AMIR^CF^ (DFG-RIsources N° RI_00052) for support in MR-imaging.

The authors thank the lighthouse Core Facility of the Medical Faculty, University of Freiburg (Project Numbers 2023/A2-Fol; 2021/B3-Fol), the DKTK, and the DFG (Project Number 450392965).

The authors express our gratitude for the China Scholarship Council’s assistance.

## Grant support and assistance

This study was funded by the German Cancer Aid, Deutsche Krebshilfe, Project ID 70113697 (to D.A.R.) and by the German Research Foundation, Deutsche Forschungsgemeinschaft (DFG), CRC1479 (Project ID 441891347, P17 to D.A.R., S01 to M.B., S02 to W.R.). M.B. is supported by the DFG, within CRC1160 (Project ID 256073931, Z02), CRC/TRR167 (Project ID 259373024, Z01), CRC1453 (Project ID 431984000, S01), TRR 359 (Project ID 491676693, Z01), TRR 353 (Project ID 471011418-SP02) and FOR 5476 UcarE (Project ID 493802833, P07). M.B. also acknowledges funding from the German Federal Ministry of Education and Research (BMBF) within the National Decade against Cancer program for PM4Onco–FKZ 01ZZ2322A and SATURN3 (01KD2206L) and G.A. and T.D. is supported by the BMBF within the Medical Informatics Funding Scheme via EkoEstMed–FKZ 01ZZ2015. XC is supported by the China Scholarship Council (Project ID 202208080226). T.C. is supported by National Natural Science Foundation of China (Project ID 82400781).

## Competing interests

The authors declare no competing interests.

## Data availability

RNAseq, scRNAseq and all other data supporting the findings of this study are available from the corresponding author upon reasonable request.

## Notes

### Competing Interest Statement

The authors have declared no competing interest.

### Author Declarations

All patients provided written informed consent prior to surgery, and tissue collection was in accordance with the Declaration of Helsinki and approved by the institutional Ethics Committee of the University of Freiburg (No. 126/17 and No. 73/18).

## References

1 Rahib, L., Wehner, M. R., Matrisian, L. M. & Nead, K. T. Estimated Projection of US Cancer Incidence and Death to 2040. JAMA Netw Open 4, e214708, doi:10.1001/jamanetworkopen.2021.4708 (2021).

2 Halbrook, C. J., Lyssiotis, C. A., Pasca di Magliano, M. & Maitra, A. Pancreatic cancer: Advances and challenges. Cell 186, 1729–1754, doi:10.1016/j.cell.2023.02.014 (2023).

3 Siegel, R. L., Giaquinto, A. N. & Jemal, A. Cancer statistics, 2024. CA Cancer J Clin 74, 12–49, doi:10.3322/caac.21820 (2024).

4 Ruess, D. A., Gorgulu, K., Wormann, S. M. & Algul, H. Pharmacotherapeutic Management of Pancreatic Ductal Adenocarcinoma: Current and Emerging Concepts. Drugs Aging 34, 331–357, doi:10.1007/s40266-017-0453-y (2017).

5 Rojas, L. A. et al. Personalized RNA neoantigen vaccines stimulate T cells in pancreatic cancer. Nature 618, 144–150, doi:10.1038/s41586-023-06063-y (2023).

6 Wasko, U. N. et al. Tumour-selective activity of RAS-GTP inhibition in pancreatic cancer. Nature 629, 927–936, doi:10.1038/s41586-024-07379-z (2024).

7 Kemp, S. B. et al. Efficacy of a Small-Molecule Inhibitor of KrasG12D in Immunocompetent Models of Pancreatic Cancer. Cancer Discov 13, 298–311, doi:10.1158/2159-8290.CD-22-1066 (2023).

8 Cancer Genome Atlas Research Network. Electronic address, a. a. d. h. e. & Cancer Genome Atlas Research, N. Integrated Genomic Characterization of Pancreatic Ductal Adenocarcinoma. Cancer Cell 32, 185–203 e113, doi:10.1016/j.ccell.2017.07.007 (2017).

9 Moore, A. R., Rosenberg, S. C., McCormick, F. & Malek, S. RAS-targeted therapies: is the undruggable drugged? Nat Rev Drug Discov 19, 533–552, doi:10.1038/s41573-020-0068-6 (2020).

10 Bailey, P. et al. Genomic analyses identify molecular subtypes of pancreatic cancer. Nature 531, 47–52, doi:10.1038/nature16965 (2016).

11 Chan-Seng-Yue, M. et al. Transcription phenotypes of pancreatic cancer are driven by genomic events during tumor evolution. Nat Genet 52, 231–240, doi:10.1038/s41588-019-0566-9 (2020).

12 Collisson, E. A., Bailey, P., Chang, D. K. & Biankin, A. V. Molecular subtypes of pancreatic cancer. Nat Rev Gastroenterol Hepatol 16, 207–220, doi:10.1038/s41575-019-0109-y (2019).

13 Moffitt, R. A. et al. Virtual microdissection identifies distinct tumor– and stroma-specific subtypes of pancreatic ductal adenocarcinoma. Nat Genet 47, 1168–1178, doi:10.1038/ng.3398 (2015).

14 Puleo, F. et al. Stratification of Pancreatic Ductal Adenocarcinomas Based on Tumor and Microenvironment Features. Gastroenterology 155, 1999–2013 e1993, doi:10.1053/j.gastro.2018.08.033 (2018).

15 Collisson, E. A. et al. Subtypes of pancreatic ductal adenocarcinoma and their differing responses to therapy. Nat Med 17, 500–503, doi:10.1038/nm.2344 (2011).

16 Singh, A. et al. A gene expression signature associated with “K-Ras addiction” reveals regulators of EMT and tumor cell survival. Cancer Cell 15, 489–500, doi:10.1016/j.ccr.2009.03.022 (2009).

17 Mueller, S. et al. Evolutionary routes and KRAS dosage define pancreatic cancer phenotypes. Nature 554, 62–68, doi:10.1038/nature25459 (2018).

18 Miyabayashi, K. et al. Intraductal Transplantation Models of Human Pancreatic Ductal Adenocarcinoma Reveal Progressive Transition of Molecular Subtypes. Cancer Discov 10, 1566–1589, doi:10.1158/2159-8290.CD-20-0133 (2020).

19 Zhao, Y. et al. Diverse alterations associated with resistance to KRAS(G12C) inhibition. Nature 599, 679–683, doi:10.1038/s41586-021-04065-2 (2021).

20 Awad, M. M. et al. Acquired Resistance to KRAS(G12C) Inhibition in Cancer. N Engl J Med 384, 2382–2393, doi:10.1056/NEJMoa2105281 (2021).

21 Dilly, J. et al. Mechanisms of Resistance to Oncogenic KRAS Inhibition in Pancreatic Cancer. Cancer Discov 14, 2135–2161, doi:10.1158/2159-8290.CD-24-0177 (2024).

22 Ruess, D. A. et al. Mutant KRAS-driven cancers depend on PTPN11/SHP2 phosphatase. Nat Med 24, 954–960, doi:10.1038/s41591-018-0024-8 (2018).

23 Fedele, C. et al. SHP2 Inhibition Prevents Adaptive Resistance to MEK Inhibitors in Multiple Cancer Models. Cancer Discov 8, 1237–1249, doi:10.1158/2159-8290.CD-18-0444 (2018).

24 Drilon, A. et al. SHP2 Inhibition Sensitizes Diverse Oncogene-Addicted Solid Tumors to Re-treatment with Targeted Therapy. Cancer Discov 13, 1789–1801, doi:10.1158/2159-8290.CD-23-0361 (2023).

25 Frank, K. J. et al. Extensive preclinical validation of combined RMC-4550 and LY3214996 supports clinical investigation for KRAS mutant pancreatic cancer. Cell Rep Med 3, 100815, doi:10.1016/j.xcrm.2022.100815 (2022).

26 Chen, X. et al. Tyrosine phosphatase PTPN11/SHP2 in solid tumors – bull’s eye for targeted therapy? Front Immunol 15, 1340726, doi:10.3389/fimmu.2024.1340726 (2024).

27 Prahallad, A. et al. CRISPR Screening Identifies Mechanisms of Resistance to KRASG12C and SHP2 Inhibitor Combinations in Non-Small Cell Lung Cancer. Cancer Res 83, 4130–4141, doi:10.1158/0008-5472.CAN-23-1127 (2023).

28 Barkley, D. et al. Cancer cell states recur across tumor types and form specific interactions with the tumor microenvironment. Nat Genet 54, 1192–1201, doi:10.1038/s41588-022-01141-9 (2022).

29 Franca, G. S. et al. Cellular adaptation to cancer therapy along a resistance continuum. Nature 631, 876–883, doi:10.1038/s41586-024-07690-9 (2024).

30 Hwang, W. L. et al. Single-nucleus and spatial transcriptome profiling of pancreatic cancer identifies multicellular dynamics associated with neoadjuvant treatment. Nat Genet 54, 1178–1191, doi:10.1038/s41588-022-01134-8 (2022).

31 Tiriac, H. et al. Organoid Profiling Identifies Common Responders to Chemotherapy in Pancreatic Cancer. Cancer Discov 8, 1112–1129, doi:10.1158/2159-8290.CD-18-0349 (2018).

32 Raghavan, S. et al. Microenvironment drives cell state, plasticity, and drug response in pancreatic cancer. Cell 184, 6119–6137 e6126, doi:10.1016/j.cell.2021.11.017 (2021).

33 Klomp, J. A. et al. Defining the KRAS– and ERK-dependent transcriptome in KRAS-mutant cancers. Science 384, eadk0775, doi:10.1126/science.adk0775 (2024).

34 Debaugnies, M. et al. RHOJ controls EMT-associated resistance to chemotherapy. Nature 616, 168–175, doi:10.1038/s41586-023-05838-7 (2023).

35 Goyal, Y. et al. Diverse clonal fates emerge upon drug treatment of homogeneous cancer cells. Nature 620, 651–659, doi:10.1038/s41586-023-06342-8 (2023).

36 Shibue, T. & Weinberg, R. A. EMT, CSCs, and drug resistance: the mechanistic link and clinical implications. Nat Rev Clin Oncol 14, 611–629, doi:10.1038/nrclinonc.2017.44 (2017).

37 Boumahdi, S. & de Sauvage, F. J. The great escape: tumour cell plasticity in resistance to targeted therapy. Nat Rev Drug Discov 19, 39–56, doi:10.1038/s41573-019-0044-1 (2020).

38 Adachi, Y. et al. Epithelial-to-Mesenchymal Transition is a Cause of Both Intrinsic and Acquired Resistance to KRAS G12C Inhibitor in KRAS G12C-Mutant Non-Small Cell Lung Cancer. Clin Cancer Res 26, 5962–5973, doi:10.1158/1078-0432.CCR-20-2077 (2020).

39 Ning, W., Marti, T. M., Dorn, P. & Peng, R. W. Non-genetic adaptive resistance to KRAS(G12C) inhibition: EMT is not the only culprit. Front Oncol 12, 1004669, doi:10.3389/fonc.2022.1004669 (2022).

40 Edwards, A. C. et al. TEAD Inhibition Overcomes YAP1/TAZ-Driven Primary and Acquired Resistance to KRASG12C Inhibitors. Cancer Res 83, 4112–4129, doi:10.1158/0008-5472.CAN-23-2994 (2023).

41 Niederst, M. J. & Benes, C. H. EMT twists the road to PI3K. Cancer Discov 4, 149–151, doi:10.1158/2159-8290.CD-13-1030 (2014).

42 Kim, D. et al. Pan-KRAS inhibitor disables oncogenic signalling and tumour growth. Nature 619, 160–166, doi:10.1038/s41586-023-06123-3 (2023).

43 Randriamanantsoa, S. et al. Spatiotemporal dynamics of self-organized branching in pancreas-derived organoids. Nat Commun 13, 5219, doi:10.1038/s41467-022-32806-y (2022).

44 Viswanathan, V. S. et al. Dependency of a therapy-resistant state of cancer cells on a lipid peroxidase pathway. Nature 547, 453–457, doi:10.1038/nature23007 (2017).

45 Schwab, A. et al. Zeb1 mediates EMT/plasticity-associated ferroptosis sensitivity in cancer cells by regulating lipogenic enzyme expression and phospholipid composition. Nat Cell Biol 26, 1470–1481, doi:10.1038/s41556-024-01464-1 (2024).

46 Genovese, G. et al. Synthetic vulnerabilities of mesenchymal subpopulations in pancreatic cancer. Nature 542, 362–366, doi:10.1038/nature21064 (2017).

47 Bolger, A. M., Lohse, M. & Usadel, B. Trimmomatic: a flexible trimmer for Illumina sequence data. Bioinformatics 30, 2114–2120, doi:10.1093/bioinformatics/btu170 (2014).

48 Dobin, A. et al. STAR: ultrafast universal RNA-seq aligner. Bioinformatics 29, 15–21, doi:10.1093/bioinformatics/bts635 (2013).

49 Liberzon, A. et al. The Molecular Signatures Database (MSigDB) hallmark gene set collection. Cell Syst 1, 417–425, doi:10.1016/j.cels.2015.12.004 (2015).

50. Korotkevich, G., et al. Fast gene set enrichment analysis. bioRxiv, 060012, doi:10.1101/060012 (2021).

51 Ritchie, M. E. et al. limma powers differential expression analyses for RNA-sequencing and microarray studies. Nucleic Acids Res 43, e47, doi:10.1093/nar/gkv007 (2015).

52 Yu, G., Wang, L. G., Han, Y. & He, Q. Y. clusterProfiler: an R package for comparing biological themes among gene clusters. OMICS 16, 284–287, doi:10.1089/omi.2011.0118 (2012).

53 Luo, W. & Brouwer, C. Pathview: an R/Bioconductor package for pathway-based data integration and visualization. Bioinformatics 29, 1830–1831, doi:10.1093/bioinformatics/btt285 (2013).

54 Wu, T. et al. clusterProfiler 4.0: A universal enrichment tool for interpreting omics data. Innovation (Camb) 2, 100141, doi:10.1016/j.xinn.2021.100141 (2021).

55 Wolf, F. A., Angerer, P. & Theis, F. J. SCANPY: large-scale single-cell gene expression data analysis. Genome Biol 19, 15, doi:10.1186/s13059-017-1382-0 (2018).

56 Satija, R., Farrell, J. A., Gennert, D., Schier, A. F. & Regev, A. Spatial reconstruction of single-cell gene expression data. Nat Biotechnol 33, 495–502, doi:10.1038/nbt.3192 (2015).

57 Korsunsky, I. et al. Fast, sensitive and accurate integration of single-cell data with Harmony. Nat Methods 16, 1289–1296, doi:10.1038/s41592-019-0619-0 (2019).

58 Lim, H. S. & Qiu, P. Quantifying Cell-Type-Specific Differences of Single-Cell Datasets Using Uniform Manifold Approximation and Projection for Dimension Reduction and Shapley Additive exPlanations. J Comput Biol 30, 738–750, doi:10.1089/cmb.2022.0366 (2023).

59 Traag, V. A., Waltman, L. & van Eck, N. J. From Louvain to Leiden: guaranteeing well-connected communities. Sci Rep 9, 5233, doi:10.1038/s41598-019-41695-z (2019).

60 Xu, S. et al. Using clusterProfiler to characterize multiomics data. Nat Protoc 19, 3292–3320, doi:10.1038/s41596-024-01020-z (2024).

61 Boj, S. F. et al. Organoid models of human and mouse ductal pancreatic cancer. Cell 160, 324–338, doi:10.1016/j.cell.2014.12.021 (2015).

