## Supplementary Figures. S1 to S15 for "Adaptive resistance to SHP2-based vertical RAS-pathway inhibition in pancreatic cancer involves multifaceted routes towards dedifferentiation"

**This PDF file includes:**

Figures. S1 to S15

### Supplementary Figure 1

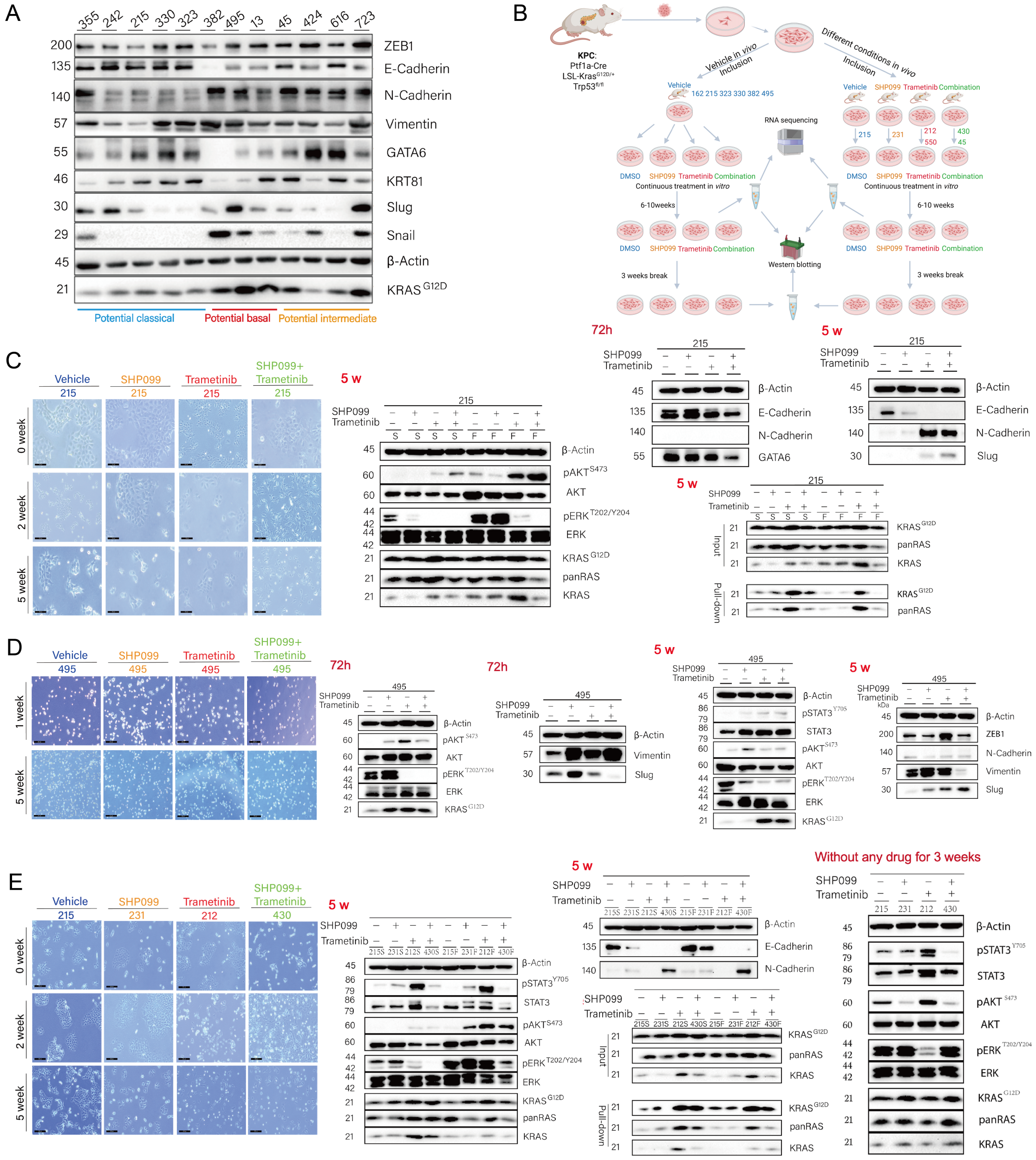

**Phenotypic and molecular adaptation of KPC-derived PDAC cell lines to SHP2-, MEK- and dual SHP2/MEK-inhibition**

(**A**) Western Blot analysis of murine KPC cell lines for PDAC subtype related proteins. β-Actin served as the loading control. (**B**) Schematic overview of the experimental design for 2D KPC cell line treatment: KPC-derived PDAC cell lines were subjected to various treatment conditions *in vitro*, including SHP2 inhibitor (SHP099), MEK inhibitor (Trametinib), and SHP099+Trametinib, short term (72h), followed by continuous treatment for 5-10 weeks to induce adaptice resistance, followed by treatment interruption for an additional 3 weeks. Samples were harvested for RNA-seq and protein analysis at defined timepoints as indicated. (**C+D**) Light microscopic appearance and immunoblot assays with the indicated antibodies of the short-term treated (72 h) and drug-resistant (5 w) KPC "classical" cell line 215 in C and KPC "basal-like" cell line 495 in D. Scale bars: 200 µm. β-Actin served as the loading control. S: FBS starvation the day before lysing the cells; F: FBS starvation the day before + supplying FBS 10 minutes before lysing the cells. Pulldown of RAS-GTP was achieved using RAF-RBD agarose beads. (**E**) Light microscopic appearance and immunoblot assays with the indicated antibodies of drug-resistant (5 w) cell lines derived from KPC mice that were given different treatments in *vivo* and then continued treatment in *vitro*. Immunoblot assays with the indicated antibodies of withdrawal of treatment for three weeks after the above four cell lines developed drug resistance. Scale bars:200 µm. β-Actin served as the loading control.

### Supplementary Figure 2

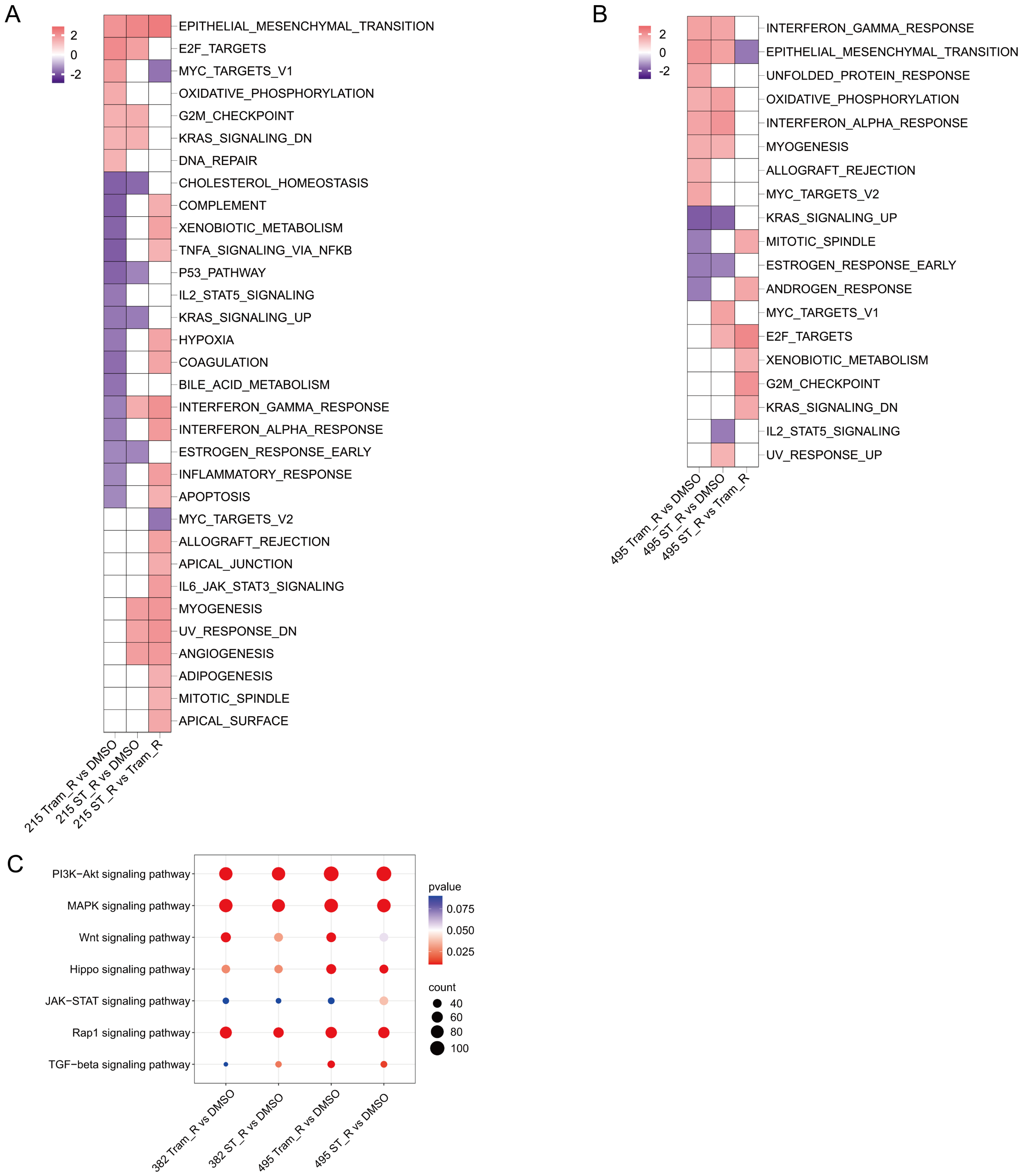

**Transcriptome analysis of KPC-derived PDAC cell lines of different subtypes with adaptive resistance to MEK and dual SHP2/MEK-inhibition**

(**A**) GSEA using the Hallmark gene set collection comparing KPC cell lines derived from parental line 215 “classical” resistant to Trametinib (215 Tram_R) and SHP099 + Trametinib (215 ST_R) with a DMSO-treated control. Normalized enrichment scores (NES) are color coded. Only gene sets with adj. $p < 0.05$ are indicated. (**B**) NES from the Hallmark gene sets comparing KPC cell lines derived from parental line 495 “basal-like” resistant to Trametinib (495 Tram_R) and SHP099 + Trametinib (495 ST_R) with a DMSO-treated control. Only gene sets with adj. $p < 0.05$ are indicated. (**C**) Pathway analysis of key signaling pathways altered in resistant PDAC cell lines (from parental 382 and 495, “basal-like”). Dot size indicates gene count; color reflects statistical significance (adj. p-value).

#### Supplementary Figure 3

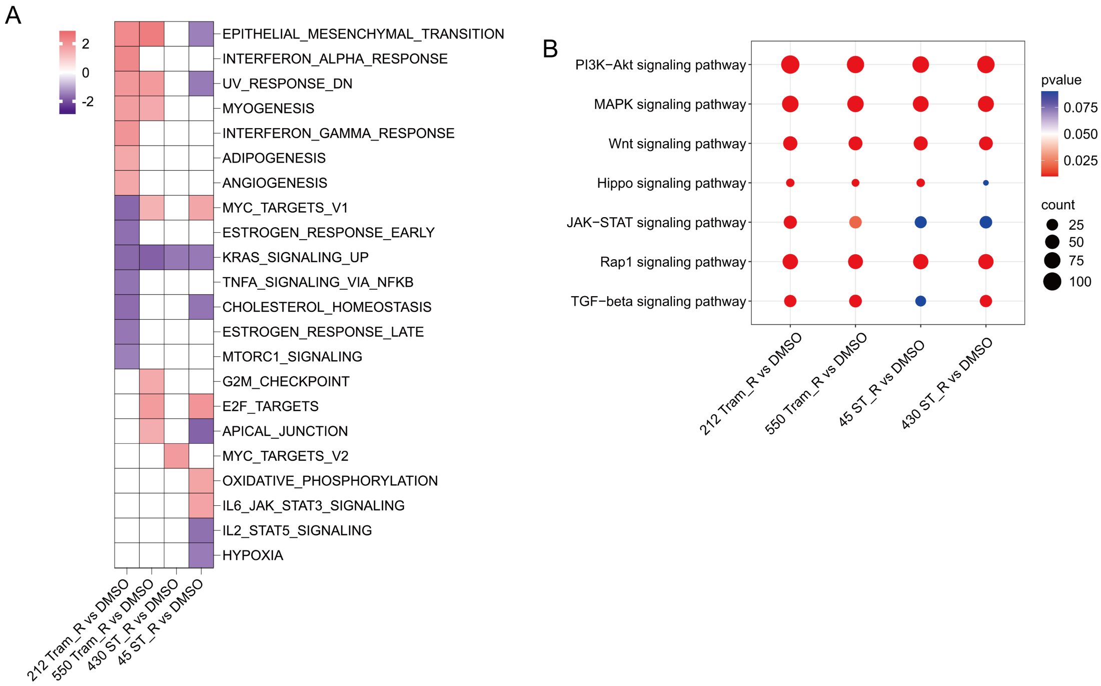

**Transcriptome analysis of KPC-derived PDAC cell lines with continued treatment with MEK and dual SHP2/MEK-inhibition *ex vivo in vitro***

(**A**) GSEA using the Hallmark gene set collection comparing KPC cell lines established from long-term *in vivo* treated PDAC tumors with *in vitro* continued treatment (_R) in comparison to *in vitro* treatment discontinuation (DMSO). Two cell lines from long-term Trametinib treated mice (212 Tram_R and 550 Tram_R, each in comparison to its DMSO counterpart) and two cell lines from long-term SHP099 + Trametinib treated mice (430 ST_R and 45 ST_R, each in comparison to its DMSO counterpart) were analyzed. Normalized enrichment scores are color coded. Only gene sets with adj. $p < 0.05$ are indicated. (**B**) Pathway analysis of key signaling pathways altered in the same cell lines as in (A). Dot size indicates gene count; color reflects statistical significance (adj. p-value).

### Supplementary Figure 4

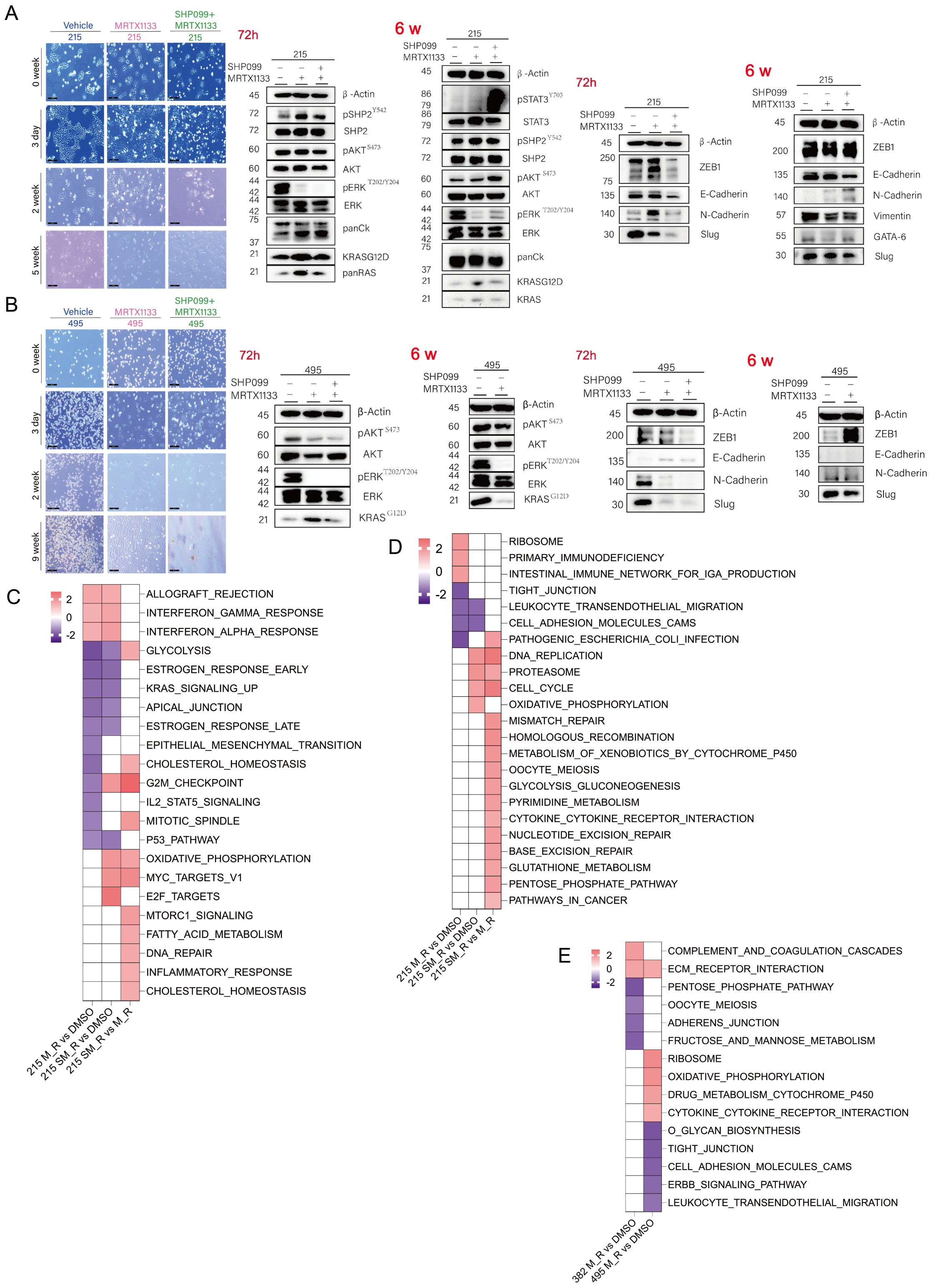

**Phenotypic and molecular adaptation of KPC-derived PDAC cell lines with adaptive resistance to $KRAS^{G12D}$ - and dual SHP2/ $KRAS^{G12D}$ -inhibition**

(**A+B**) Light microscopic appearance and immunoblot assays with the indicated antibodies of short-term treated (72 h) and drug-resistant KPC "classical" cell line 215 (6 weeks of treatment) in (A) and KPC "basal-like" cell line 495 (9 weeks of treatment) in (B). Scale bars: 200 µm. β-Actin served as the loading control. (**C+D**) GSEA using the (C) Hallmark and (D) KEGG gene sets comparing KPC cell lines derived from parental line 215 "classical" resistant to MRTX1133 (215 M_R) and SHP099 + MRTX1133 (215 SM_R) with a DMSO-treated control. Normalized enrichment scores are color coded. Only gene sets with adj. $p < 0.05$ are indicated. (**E**) GSEA using KEGG gene sets comparing KPC cell lines derived from parental lines 382 and 495 "basal-like" resistant to MRTX1133 (382 M_R and 495 M_R) with a DMSO-treated control. Normalized enrichment scores are color coded. Only gene sets with adj. $p < 0.05$ are indicated.

### Supplementary Figure 5

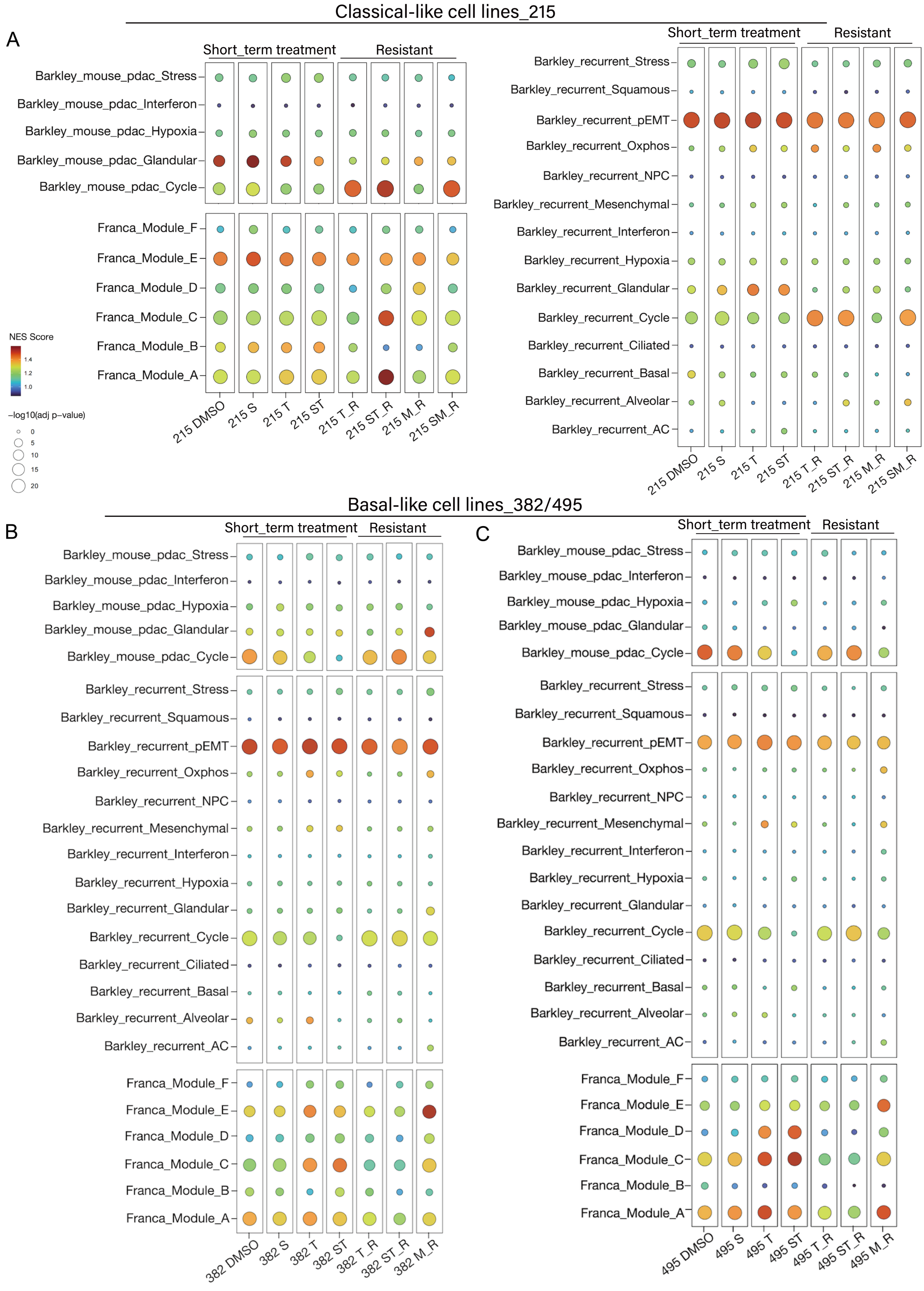

**Enrichment analysis for cancer cell states and resistance cell state transitions in KPC cell lines**

(**A**) Single sample enrichment analysis (ssGSEA) using the Barkley *et al.* gene modules, the catalog of gene modules whose expression defines recurrent cancer cell states described by Barkley *et al.* and the gene modules described by Franca *et al.,* representing a resistance continuum early cell state of adaptive resistance (gene module A) to the latest stage of adaptive resistance (gene module F). Dot plot illustrating enrichment of modules A to F across multiple treatment conditions (Short_term treatment: 72 h, Resistant: 5 w or 6 w) in the "classical" 215 cell line. NES scores and adj. p-values are visualized by color and dot size, respectively. (**B-C**) ssGSEA using the Barkley *et al.* gene modules, the catalog of gene modules whose expression defines recurrent cancer cell states described by Barkley *et al.* and the gene modules described by Franca *et al.,* representing a resistance continuum early cell state of adaptive resistance (gene module A) to the latest stage of adaptive resistance (gene module F). Dot plot illustrating enrichment of modules A to F across multiple treatment conditions (Short_term treatment: 72 h, Resistant: 9 weeks treatment) in the "basal" 382 and 495 cell lines. NES scores and adj. p-values are visualized by color and dot size, respectively.

**Supplementary Figure 6**

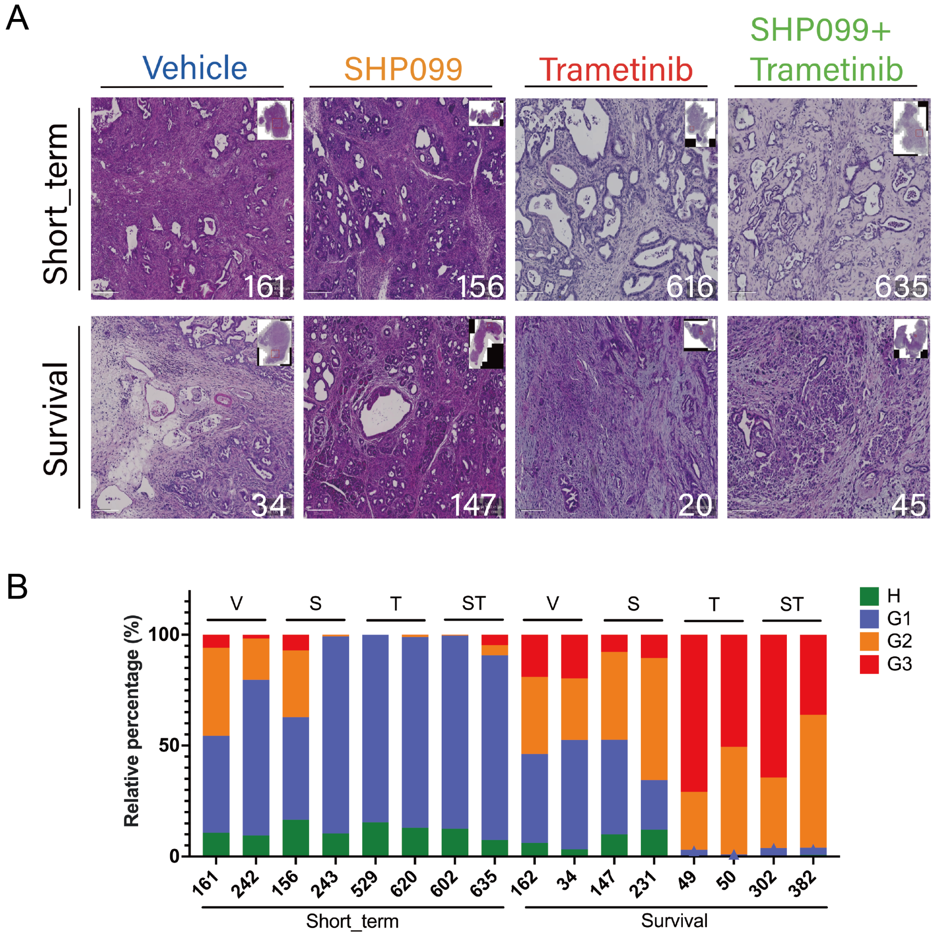

**Histological grading of pancreatic tumors from KPC mice having received different treatment regimens**

Hematoxylin and eosin (H&E) staining was performed on whole pancreatic tissues slices from KPC mice receiving short-term or survival-endpoint treatments with vehicle (V), SHP099 (S), Trametinib (T), or the combination (ST). (**A**) Representative histological images, with tumor sample IDs indicated. **(B)** One whole tumor slice was quantified per mouse for relative contribution of areas of healthy/non-transformed (H) pancreas tissue and frank PDAC with well-differentiated (G1), moderately differentiated (G2), and poorly differentiated (G3) grades. Each bar indicates one individual mouse tumor. Mouse IDs are given below.

### Supplementary Figure 7

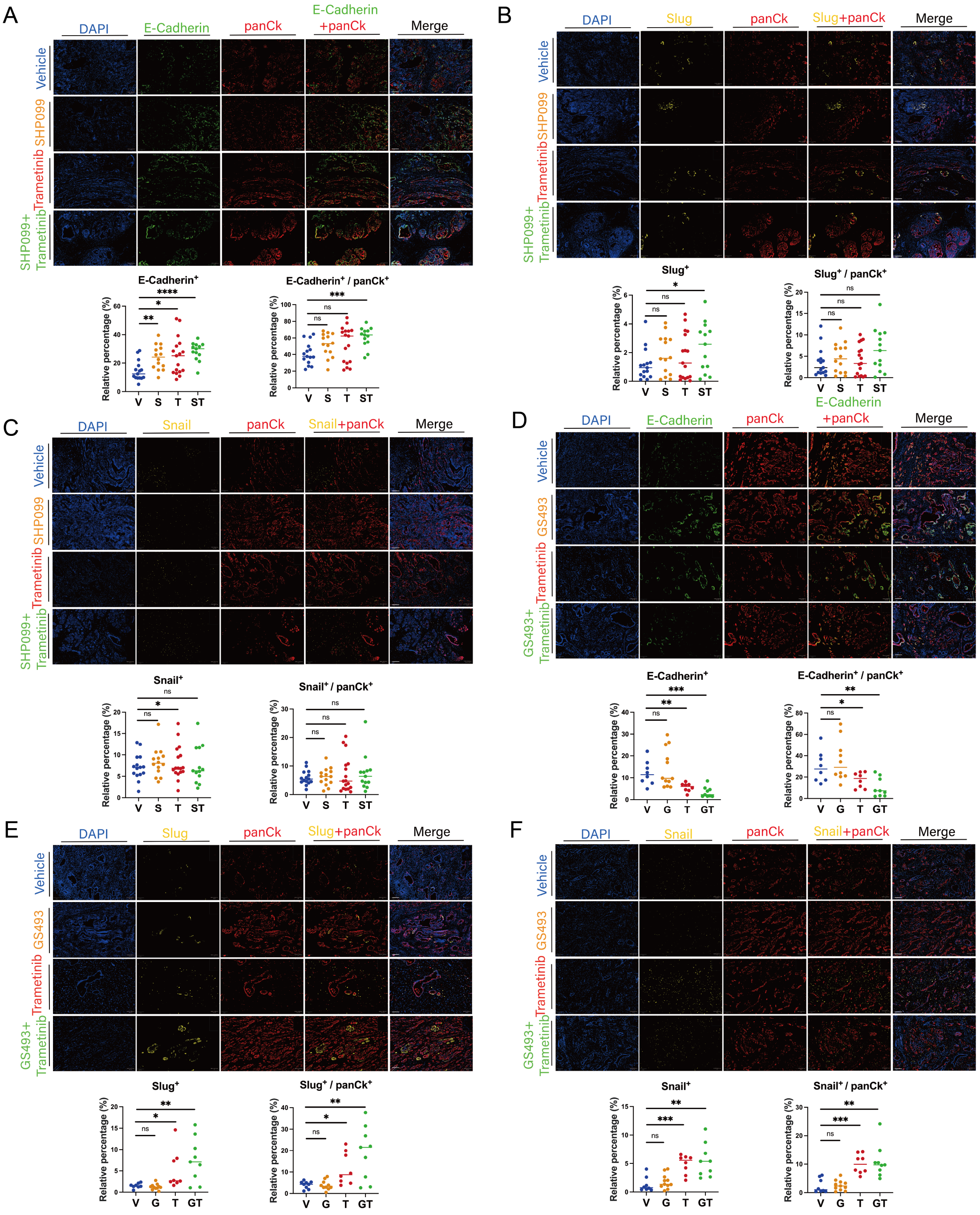

**Multiplex immunofluorescence analysis of EMT-related markers in KPC tumors following SHP2-, MEK- and dual SHP2/MEK-inhibition**

(**A-C**) **EMT-related markers in the short-term treatment cohorts (2 weeks of SHP099, Trametinib or SHP099 + Trametinib, vs. vehicle) (A**) Tumor cell specific (panCK positive) expression of E-Cadherin. V: n=15; S: n=14; T: n=17; ST: n=13. (**B**) Tumor cell specific (panCK positive) expression of Slug. V: n=15; S: n=14; T: n=17; ST: n=13. (**C**) Tumor cell specific (panCK positive) expression of Snail. V: n=15; S: n=14; T: n=17; ST: n=13. (**D-F**) **EMT accompanies evolving resistance to SHP2/MEK inhibition *in vivo*. Analysis of historical cohorts treated with GS493 and Trametinib from Ruess DA *et al*.** V: vehicle; G: GS493 (SHP2 phosphatase inhibitor); T: Trametinib (T); GT: GS493+Trametinib. (**D**) Tumor cell specific (panCK positive) expression of E-Cadherin. V: n=8; S: n=12; T: n=8; ST: n=9. (**E**) Tumor cell specific (panCK positive) expression of Slug. V: n=8; S: n=12; T: n=8; ST: n=9. (**F**) Tumor cell specific (panCK positive) expression of Snail. V: n=8; S: n=12; T: n=8; ST: n=9. Statistical significance was determined via one-way ANOVA in all panels, with comparisons made against corresponding vehicle controls. **** $p<0.0001$; *** $p<0.001$; ** $p<0.01$; * $p<0.05$; $^{ns}$ $p>0.05$.

#### Supplementary Figure 8

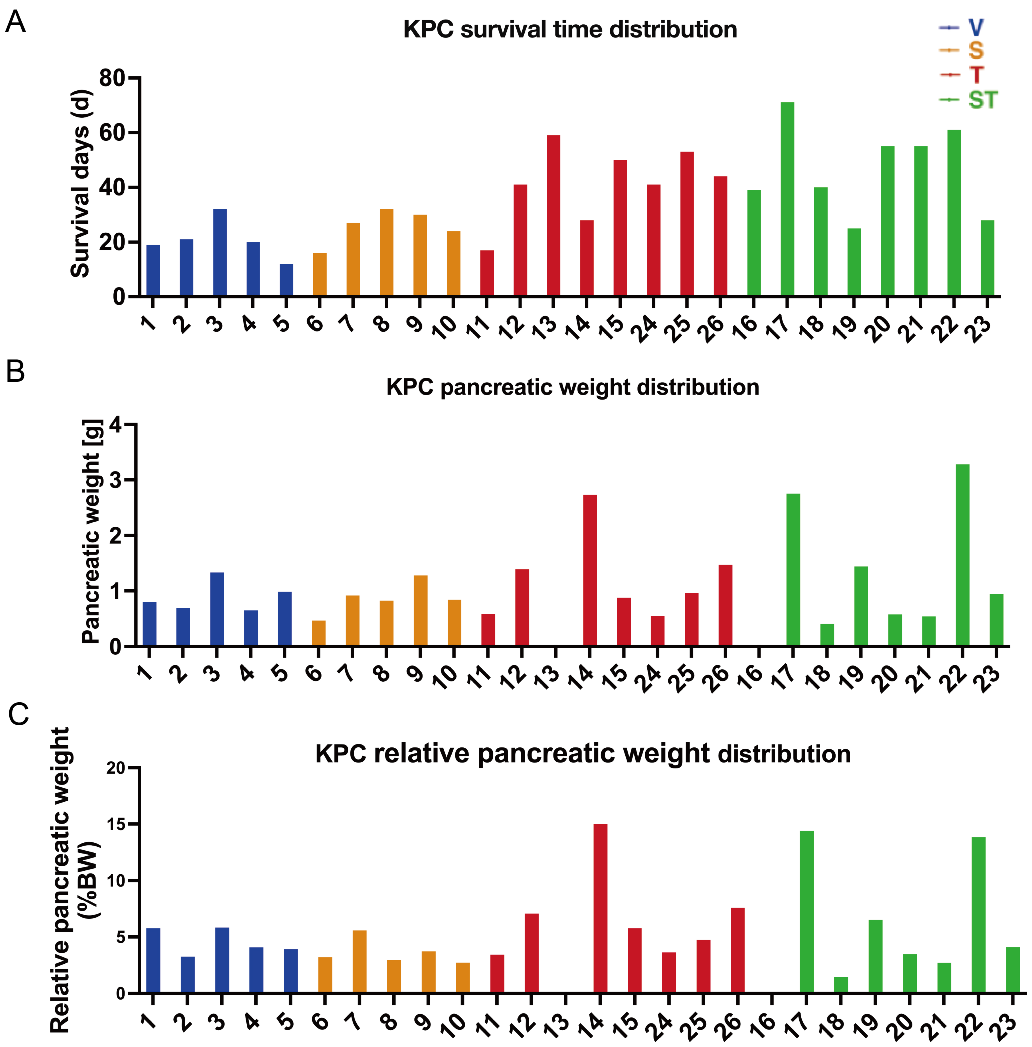

**Distribution of KPC tumor characteristics among all samples subjected to bulk tissue RNA sequencing**

(**A**) Survival time distribution (in days) of individual KPC mice included in bulk tissue RNA-seq analysis, stratified by treatment group: Vehicle (V, blue), SHP099 (S, orange), Trametinib (T, red), and SHP099+Trametinib combination (ST, green). (B) The distribution of pancreatic weight (in grams) in each KPC sample at the clinical endpoint. (**C**) Relative pancreatic weight (pancreatic weight/body weight ratio) distribution across all samples.

#### Supplementary Figure 9

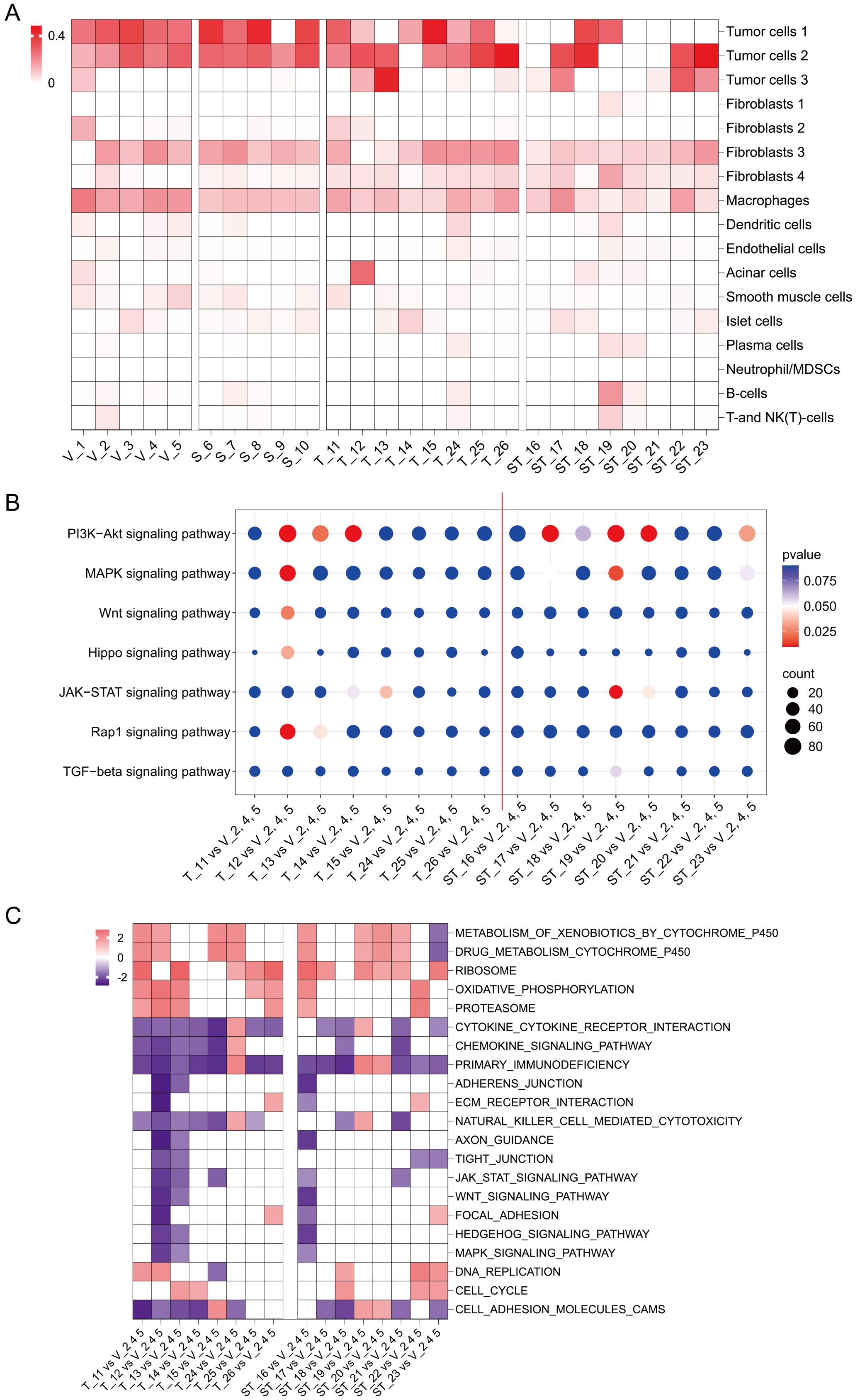

**Bulk PDAC tissue RNA sequencing deconvolution and gene set enrichment analysis of samples from the KPC survival cohort**

(**A**) *In silico* deconvolution analysis of bulk tissue transcriptomes by CIBERSORTx using the cell clusters identified in the single-cell dataset from Figure 6 and Supplenmentary 11). (**B**) Pathway analysis of key signaling pathways altered in individual long-term treated KPC mice for the comparisons of Trametinib (T) and SHP099+Trametinib (ST) treated tumors to the grouped reference (merged vehicle treated tumors 2, 4 and 5). Dot size indicates gene count; color reflects statistical significance (adj. p-value). (**C**) GSEA using the KEGG gene set collection for the same comparisons as in (A). Normalized enrichment scores are color coded. Only gene sets with adj. $p < 0.05$ are shown.

#### Supplementary Figure 10

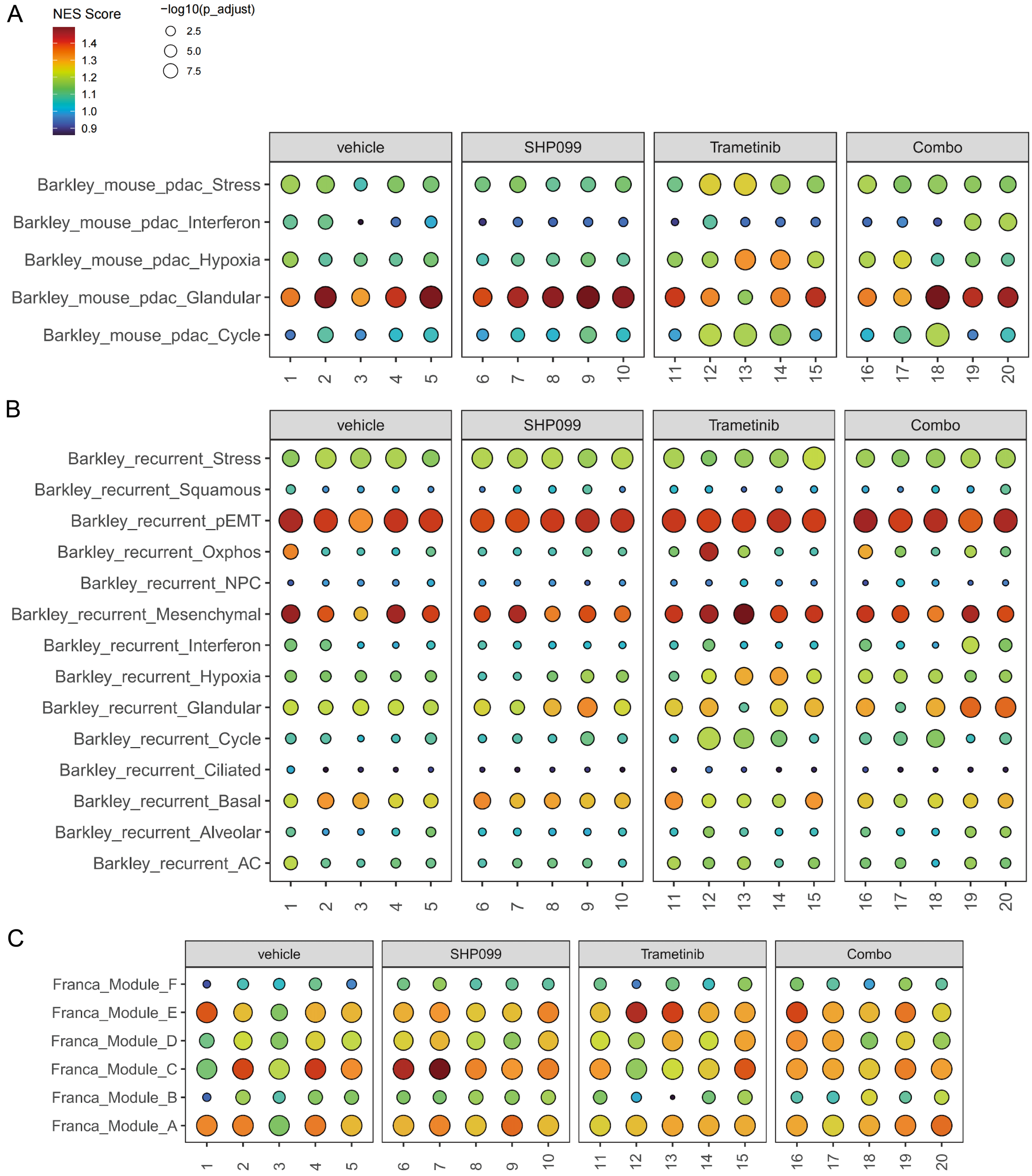

**Bulk KPC tissue RNA sequencing enrichment analysis for cancer cell states and resistance cell state transitions**

(**A-C**) Mouse PDAC cell state analysis by ssGSEA using the Barkley *et al.* gene sets in (A); ssGSEA of Barkley recurrent cancer cell state gene modules in (B); Resistance continuum gene module ssGSEA based on gene sets described by Franca *et al.* in (C). Dot plot showing enrichment of gene modules across multiple treatment conditions in individual KPC tumor tissues. NES and adj. p-values are visualized by color and dot size, respectively.

### Supplementary Figure 11

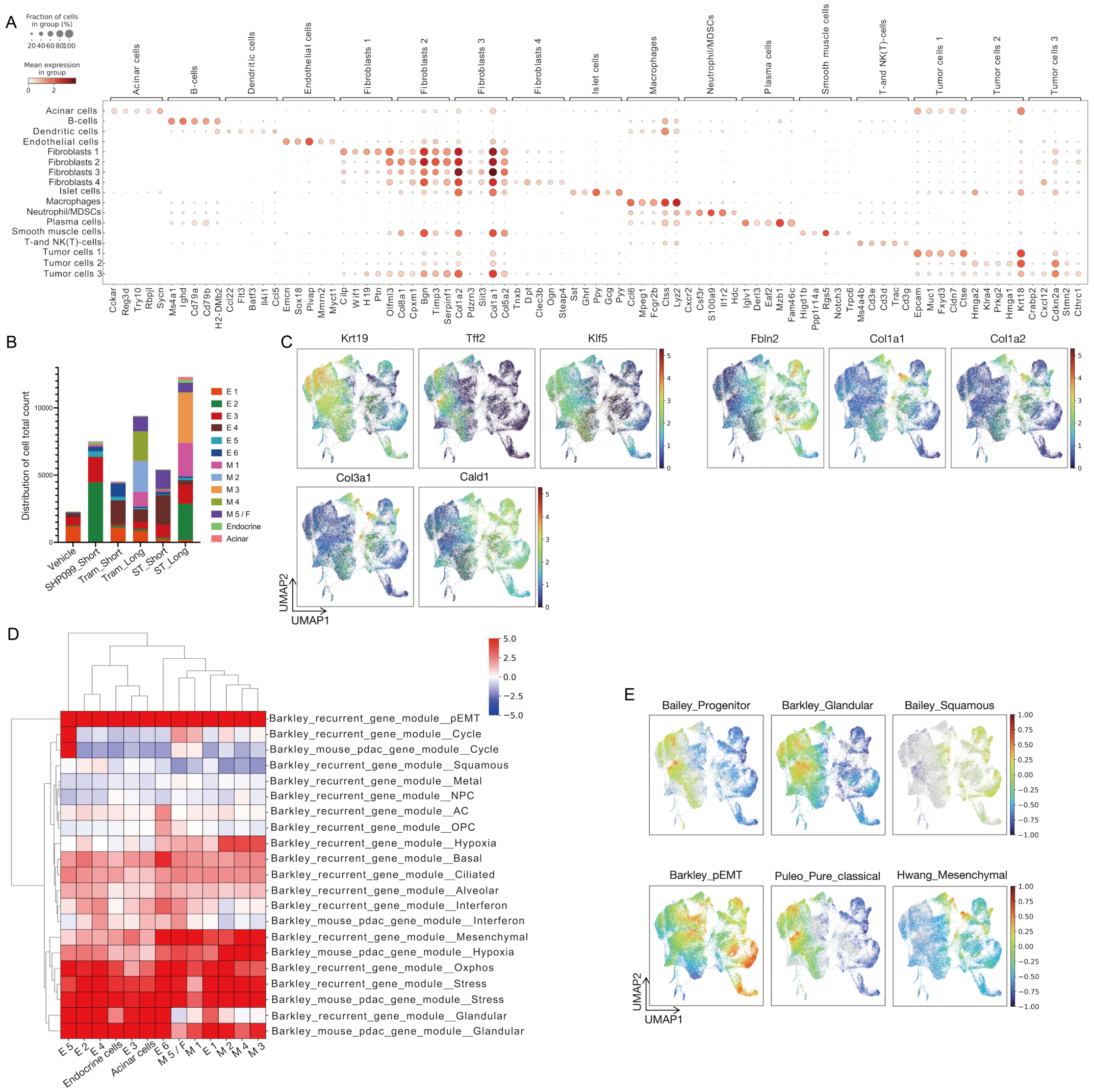

**Characterization of cell populations, marker gene expression, cell states and PDAC molecular subtypes in single-cell RNA-seq data**

(**A**) Dot plot showing the fraction and mean expression of representative marker genes across annotated 17 cell populations. Dot size indicates the fraction of cells expressing each gene, and color intensity represents the mean expression level. (**B**) Stacked bar plot showing overall populations and relative proportions of epithelial and mesenchymal cell clusters across treatment groups. “Short” indicating 2 weeks, “Long” indicating 5-6 weeks of *in vivo* treatment. Vehicle was administered for 2 weeks. (**C**) UMAP feature plots showing the expression of selected epithelial (*Krt19, Tff2, Klf5*), mesenchymal (*Fbn2, Col1a1, Col1a2, Col3a1, Cald1*) marker genes. Color intensity represents $log_2$-normalized expression levels, with higher expression shown in red and lower expression in blue. (**D**) Heatmap showing the ssGSEA enrichment scores of published recurrent cancer cell state and murine PDAC cell state gene modules (Barkley et al.) across the identified tumor/epithelial clusters (Acinar, Endocrine, E1–E6, M1–M5/F). Color scale represents normalized enrichment scores (NES). (**E**) UMAP feature plots showing gene set enrichment scores for published PDAC subtype signatures (Bailey_Progenitor, Bailey_Glandular, Bailey_Squamous, Barkley_pEMT, Puleo_Pure_classical, Hwang_Mesenchymal) across the identified tumor/epithelial clusters.

### Supplementary Figure 12

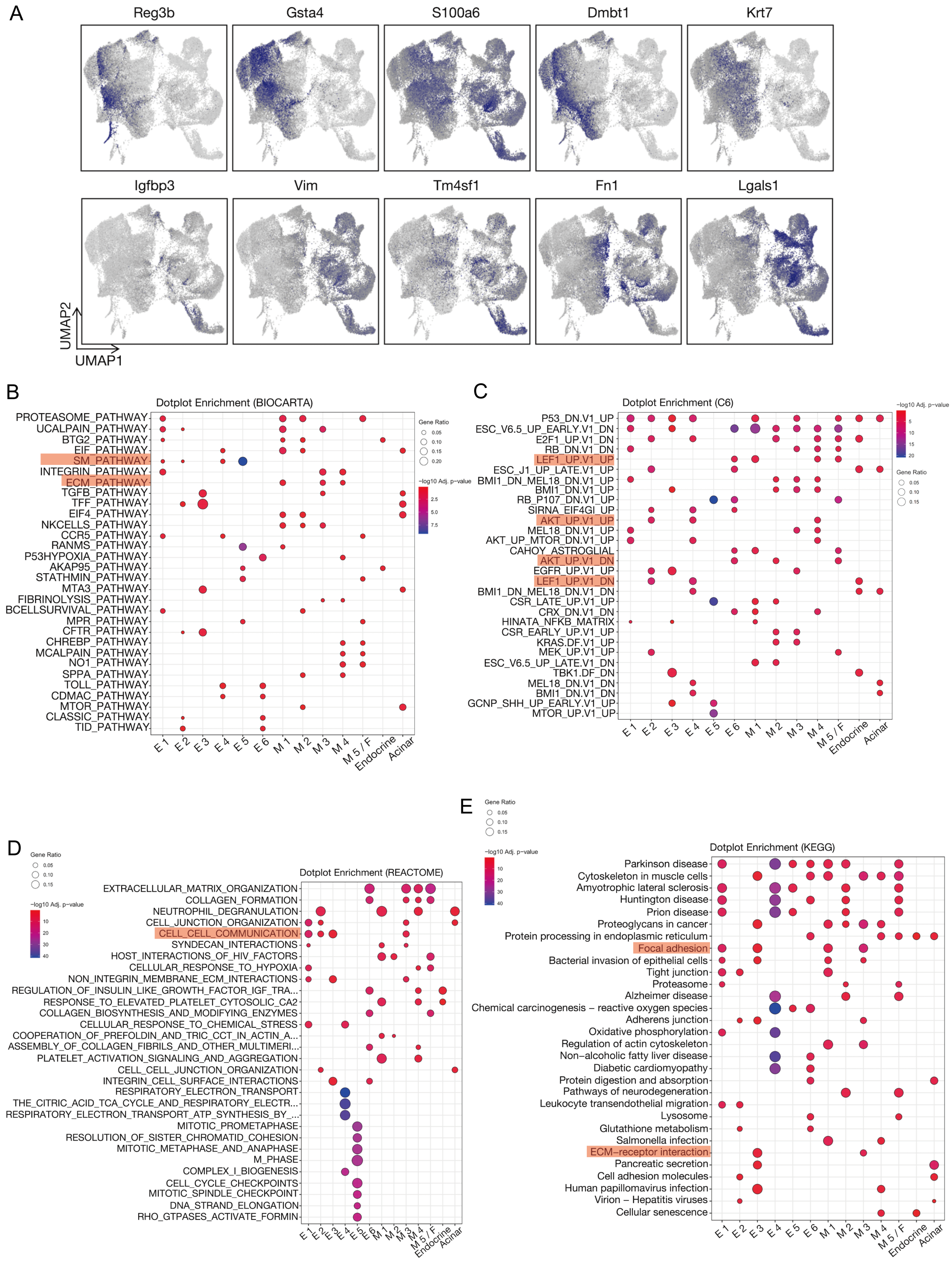

**Pseudotime-trajectory associated key gene expression and pathway enrichment analysis in the KPC scRNA seq dataset**

(**A**) UMAP feature plots showing the expression of key genes (*Reg3b, Gsta4, S100a6, Dmbt1, Krt7, Igfbp3, Vim, Tm4sf1, Fn1, Lgals1*) relevant for the pseudotime trajectory. (**B-E**) Cluster-wise GSEA. Dot plot visualization of enrichment results across identified tumor/epithelial cell clusters (E1–E6, M1–M5/F, endocrine, and acinar cells) using four gene set colletions: (**B**) BIOCARTA, (**C**) Oncogenic Signatures (MSigDB C6), (**D**) REACTOME, and (**E**) KEGG. Dot size represents the proportion of genes involved in each gene set and color indicates the –log10 adj. p value.

### Supplementary Figure 13

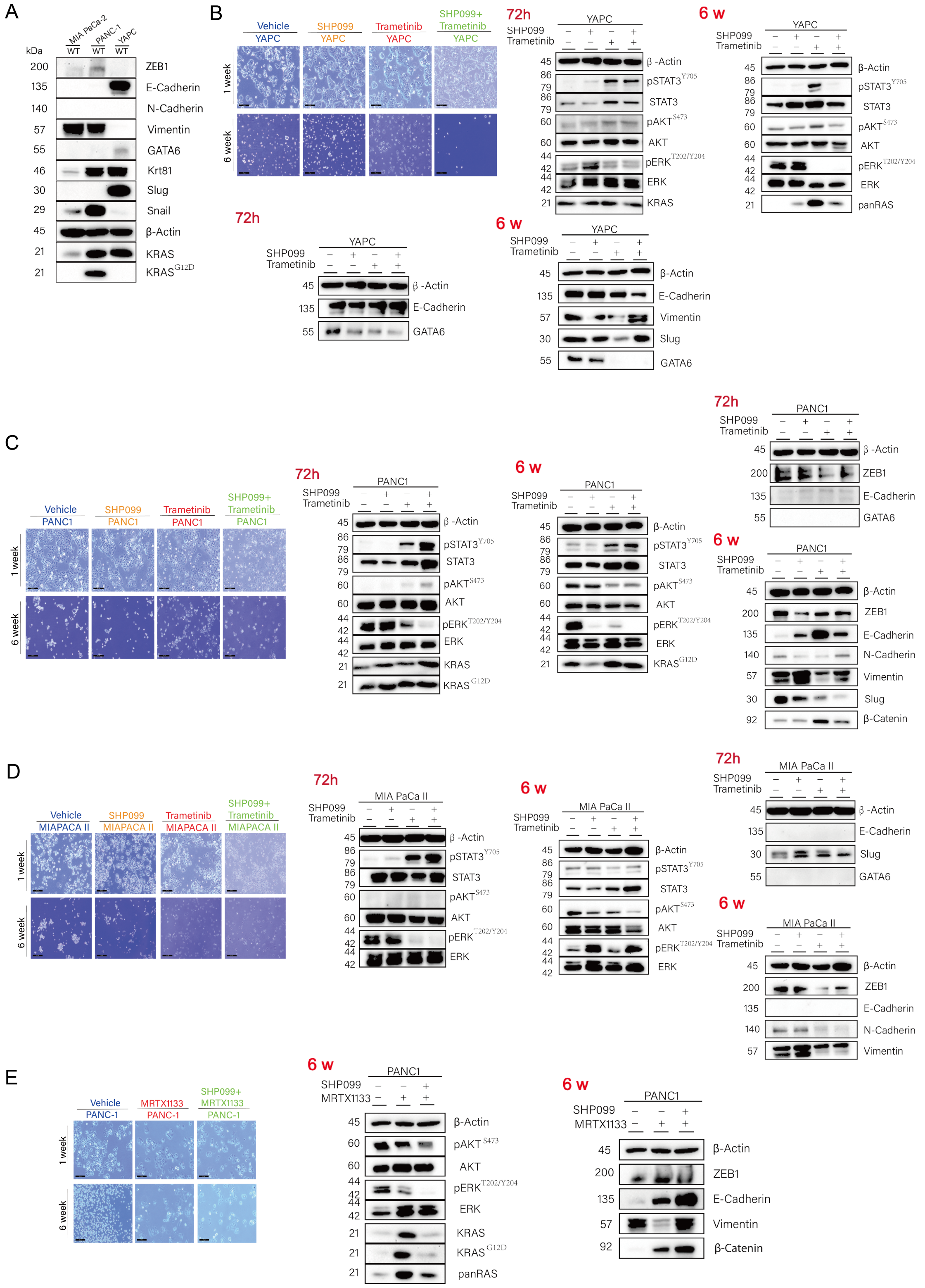

**Phenotypic and molecular adaptation of human PDAC cell lines to SHP2-, MEK- and KRAS$^{G12D}$ and dual SHP2/MEK- or dual SHP2/ KRAS$^{G12D}$ inhibition**

(**A**) Western Blot analysis of human PDAC cell lines for subtype related proteins. β-Actin served as the loading control. (B) Light microscopic appearance and immunoblot assays with the indicated antibodies of the short-term treated (72 h) and drug-resistant (6 weeks treatment) human cell line YAPC (as representative of the "classical" subtype). Scale bars: 200 µm. β-Actin served as the loading control. (**C**) Light microscopic appearance and immunoblot assays with the indicated antibodies of the short-term treated (72 h) and drug-resistant (6 weeks treatment) human cell line PANC1 (as representative of an "intermediate" subtype). Scale bars: 200 µm. β-Actin served as the loading control. (**D**) Light microscopic appearance and immunoblot assays with the indicated antibodies of the short-term treated (72 h) and drug-resistant (6 weeks treatment) human cell line MIAPACA II (as representative of the "basal-like" subtype). Scale bars: 200 µm. β-Actin served as the loading control. (**E**) Light microscopic appearance and immunoblot assays with the indicated antibodies of drug-resistant (SHP2/KRAS$^{G12D}$ inhibition, 6 weeks treatment) human cell line PANC1. Scale bars: 200 µm. β-Actin served as the loading control.

### Supplementary Figure 14

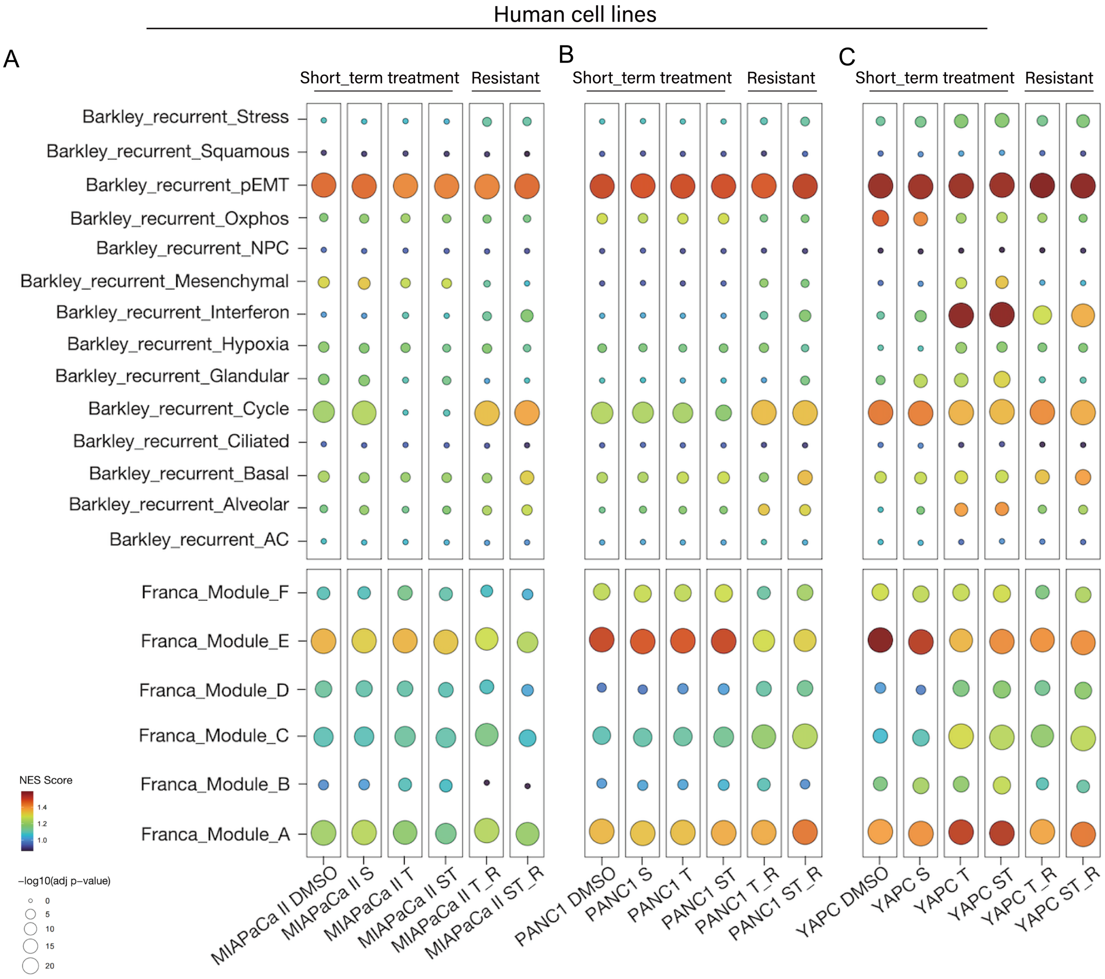

**Gene set enrichment analysis for cancer cell states and resistance cell state transitions in human PDAC cell lines**

(**A,B,C**) ssGSEA of Barkley et al. recurrent cancer cell state gene modules and resistance continuum cell state enrichment analysis based on gene modules described by Franca *et al.* for MIAPACA II , PANC1, YAPC. Dot plots showing enrichment of cell state gene modules across multiple treatment conditions (Short_term treatment: 72 h, Resistant: 6 weeks treatment). NES and adj. p-values are visualized by color and dot size, respectively.

### Supplementary Figure 15

A

| | Basal-like | | | | | | | | | | | Intermediate | | Classical-like | | | | | |
|---|---|---|---|---|---|---|---|---|---|---|---|---|---|---|---|---|---|---|---|
| | B23 | B100 | B150 | B40 | B261 | B417 | B6 | B45 | B424 | M7 | M12 | B177 | B68 | B389 | B429 | B406 | M5 | B111 | B55 |
| V | √ | √ | √ | √ | √ | √ | √ | √ | √ | √ | √ | √ | √ | √ | √ | √ | √ | √ | √ |
| T | √ | √ | √ | √ | √ | √ | √ | √ | √ | √ | √ | √ | √ | √ | √ | √ | √ | √ | √ |
| ST | – | – | √ | √ | – | – | – | – | √ | – | √ | – | √ | √ | – | – | – | – | √ |
| M | – | – | √ | √ | – | – | – | – | – | – | √ | – | √ | – | – | – | – | – | – |
| SM | – | – | – | – | – | – | – | – | – | – | – | – | – | – | – | – | – | – | – |

B

| | DMSO | Tram_R | ST_R | M_R | SM_R |
|---|---|---|---|---|---|
| M12 | ● | ○ | ○ | ○ | - |
| B150 | ● | ⊘ | ● | ⊘ | - |
| B40 | ⊘ | ⊘ | ● | ⊘ | - |
| B424 | ● | ⊘ | ⊘ | - | - |
| B100 | ⊘ | ● | ● | - | - |
| B261 | ⊘ | ⊘ | ⊘ | - | - |
| B68 | ⊘ | ⊘ | ⊘ | - | - |
| B429 | ○ | ⊘ | - | - | - |
| B389 | ○ | ⊘ | ⊘ | - | - |
| B55 | ○ | ○ | ○ | - | - |

○ mostly hollow/epithelial
⊘ mixed
● mostly solid/clustered

C

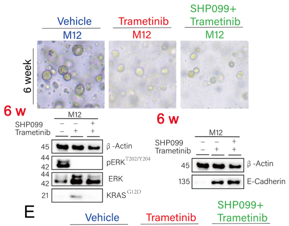


D

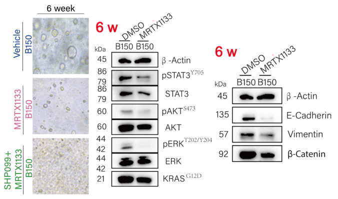


E

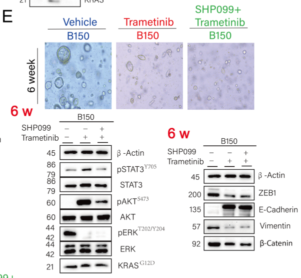


F

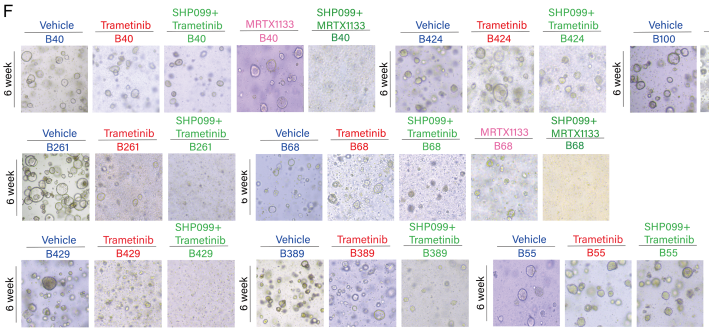


G

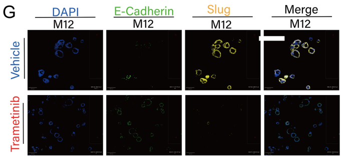

**Characterization of adaptive resistance of patient-derived PDAC organoids to MEK, KRAS$^{G12D}$ and dual SHP2/MEK and dual SHP2/KRAS$^{G12D}$ inhibition**

(**A**) Table illustrating the efficiency of generating treatment resistant PDO lines. OF the >40 PDO lines tested, for the 19 lines shown in this table an adaptive resistant line was successfully established under long-term Trametinib (T) treatment (including a DMSO vehicle control), indicated by the checkmark. PDO lines are tabulated stratified by molecular subtype of the untreated parental line (basal-like, intermediate, and classical). For 7 PDO lines organoids resistant to dual SHP099 + Trametinib (ST) were successfully established; for only 4 PDO lines organoids resistant to MRTX1133 (M) were successfully established, and it was not possible to establish organoids resistant to dual SHP099 + MRTX1133 for none of the >40 PDO lines tested. (**B**) Light microscopic morphology of PDAC organoids in untreated state (DMSO) and in treatment resistant states. Organoids were categorized based on morphology into three groups: mostly hollow/epithelial (orange circles), mostly solid/clustered (green filled circles) and mixed (red prohibition symbol). (**C-E**) Light microscopic appearance and immunoblot assays with the indicated antibodies of drug-resistant (KRAS$^{G12D}$-inhibition, 6 weeks treatment) PDO B150 (C), of drug-resistant (dual SHP2/MEK inhibition, 6 weeks treatment) PDO M12 (D) and drug-resistant (dual SHP2/MEK inhibition, 6 weeks treatment) PDO B150 (E). Scale bars: 200 µm. β-Actin served as the loading control. (**F**) Light microscopic appearance of additional PDAC organoid lines (B40, B424, B100, B261, B68, B429, B389, B55) following 6-week treatment with Trametinib, SHP099 + Trametinib, MRTX1133 and SHP099 + MRTX1133. (**G**) Representative immunofluorescence images of PDO M12 showing E-Cadherin and Slug expression following long-term treatment with vehicle or Trametinib.
